# Circulating Cell Type Senescence Signatures Reveal High-Resolution Health Status and Trajectories in Human Longitudinal Studies

**DOI:** 10.64898/2026.02.06.26345739

**Authors:** Bradley Olinger, Carlos Anerillas, Allison B. Herman, Dimitrios Tsitsipatis, Reema Banarjee, Toshiko Tanaka, Julian Candia, Manolis Maragkakis, Stefania Bandinelli, Keenan A. Walker, Eleanor M. Simonsick, Yue A Qi, Luigi Ferrucci, Myriam Gorospe, Nathan Basisty

## Abstract

Cellular senescence increases in frequency with age and is implicated in age-related pathologies, and identifying circulating biomarkers of senescence holds great diagnostic potential. Circulating senescence signatures are predictive of many age-related traits and diseases, though cell type-specific senescence signatures have not been comprehensively explored. In this study, senescence signatures from the Senescence Catalog (SenCat), including 14 human cell types such as peripheral blood mononuclear cells, renal epithelial cells, vascular smooth muscle cells, among others, are examined for their clinical relevance in circulation in two longitudinal studies: 1,275 participants of the Baltimore Longitudinal Study of Aging (BLSA) and 997 participants of the Invecchiare in Chianti (InCHIANTI) study. Notably, pooled senescence proteins outperformed non-senescence proteins in predicting many clinical parameters such as age and hypertension, and in many instances cell type senescence signatures mapped most strongly to their corresponding health domain. Importantly, the immune cell senescence signature is associated with future onset of several diseases such as diabetes. This study demonstrates that circulating cell type-specific biomarkers of senescence can reveal higher resolution health status than previously attained.

**HIGHLIGHTS:**

- Circulating senescence associated proteins tend to outperform non-senescence proteins as biomarkers of clinical phenotypes in two independent longitudinal studies.
- A core senescence signature developed from 14 human cell types predicted a range of clinical phenotypes during aging.
- Cell type senescence signatures more strongly associated with their corresponding health domains.
- The immune cell senescence signature and others were associated with mortality and diabetes onset, highlighting relevance for assessing health trajectories.

## INTRODUCTION

Cellular senescence becomes more common with age as healthy cells encounter sublethal environmental and genotoxic stress or accrue other types of damage, and is implicated in age-related decline.^1,2^ Senescence is characterized in part by proteomic expression changes, including the secretion of pro-inflammatory cytokines and other proteins, which become amplified during sustained senescence and in large part drive its deleterious effect in a chronic, age-related context.^3^ These senescence-associated proteins (SAPs) have since proven to be heterogeneous by cell type and senescence-inducing stimulus.^4^ SAPs have been characterized in a variety of tissues, including in human lung fibroblasts and epithelial cells,^5^ as well as human endothelial, epithelial, preadipocytes, fibroblasts, and myoblasts,^6^ alveolar epithelial cells,^7^ and other cell lines.^8,9^ Despite this, efforts to further characterize senescence-associated proteomic changes that are specific to each cell type (senotype), as well as those common among many cell types, are needed to better understand the heterogeneity of senescence and it’s emerging role in aging phenotypes.

One promising technique in assessing individual senescence burden is through the quantification of SAPs in circulating plasma. The plasma senescence burden has previously demonstrated compelling clinical associations, including with age,^10,11^ frailty,^6,12^ and mortality,^13^ as well as clinical parameters across four interrelated health domains, including neurodegeneration, pulmonary disease, cardiometabolic disease, and musculoskeletal disorders. Specifically, plasma senescence markers are associated with impaired mobility,^14^ poor cognition,^15–17^ neurodegenerative disease,^18,19^ impaired lung function,^20^ idiopathic pulmonary fibrosis,^21^ and diabetes,^22^ among other negative health outcomes.^23^ In recent years, a group of senescence-targeting compounds collectively known as senotherapeutics has been investigated for their limited and context dependent senescence-attenuating effects.^24^ Senotherapeutic drugs have demonstrated an ability to lower circulating SAPs in human trials,^25^ and to partially alleviate some aging phenotypes, such as increasing cognition scores among the cognitively impaired,^26^ lowering some measures of systolic blood pressure among the hypertensive,^27^ and improving some metrics of mobility.^28^ Other lifestyle and behavioral interventions such as increasing vegetable consumption,^29^ caloric restriction,^30–32^ and physical activity^33^ have also lowered some markers of the circulating senescence burden.

A remarkable recent finding is that beyond general clinical traits such as age and mortality, organ-specific proteins can be tracked in circulation and used to model organ age and organ-specific clinical traits.^34^ This finding raises the captivating possibility that beyond systemic age and overall health, circulating proteomic signatures can be used to assess the status of organ-specific health and disease. Considering the previously demonstrated clinical relevance of circulating canonical senescence signatures, examining senotype-specific senescence signatures in circulation could similarly shed light on the unique clinical relevance of organ-specific senescence. This study aims to conduct such an investigation of senescence markers derived from 14 cell types in the Senescence Catalog (SenCat) in order to determine if cell type-specific senescence proteins are more clinically relevant than non-senescence proteins and can uniquely predict and correlate with diverse and organ system-specific clinical parameters when quantified in circulating plasma in two human cohorts: The Baltimore Longitudinal Study of Aging (BLSA), and the Invecchiare in Chianti (InCHIANTI) study. Identifying cell type-specific biomarkers of senescence cultivates a better understanding of the role that senescence plays in aging and disease and provides a tool for non-invasive assessment of individual senescence burden at higher resolution than previously attained.

## RESULTS

### Identification of circulating cell type-specific senescence signatures

SAPs were collected from the SenCat, a multi-omic (transcriptomic and proteomic) catalog of senescence-associated changes in 14 primary human cell types: renal epithelial cells, small-airway epithelial cells, skeletal myoblasts, astrocytes, osteoblasts, coronary artery endothelial cells, preadipocytes, vascular smooth muscle cells, peripheral blood mononuclear cells (PBMCs), neonatal epidermal keratinocytes, neonatal epidermal melanocytes, umbilical vein endothelial cells, neonatal fibroblasts, and fetal lung fibroblasts (**Fig. 1**). SAPs were determined following multiple methods, including treatment with ionizing radiation, etoposide, or doxorubicin. Dose, incubation times, and days of incubation prior to proteomics measurement are available in **Supp. Table 1**. Given the known heterogeneity of SAPs based on senescence-induction method,^5^ cell type-specific senescence signatures were identified as those proteins that were significantly elevated in two induction methods. To compare clinical relevance between SAPs and a control group, specifically those proteins not associated with senescence in any cell type (Non-SAPs), all proteins detected in the SenCat were grouped by association with cellular senescence. Proteins were categorized as a senescence marker/signature if elevated in two induction methods for any cell type, a non-senescence marker if they were not elevated in any cell type when induced to senescence in either induction method, and inconclusive if they were elevated in a senescent cell type using only one induction method. These senescence, non-senescence, and inconclusive categories consisted of approximately 42%, 39%, and 19% of total SenCat proteins, respectively (**Fig. 2a**).

**Figure 1:**
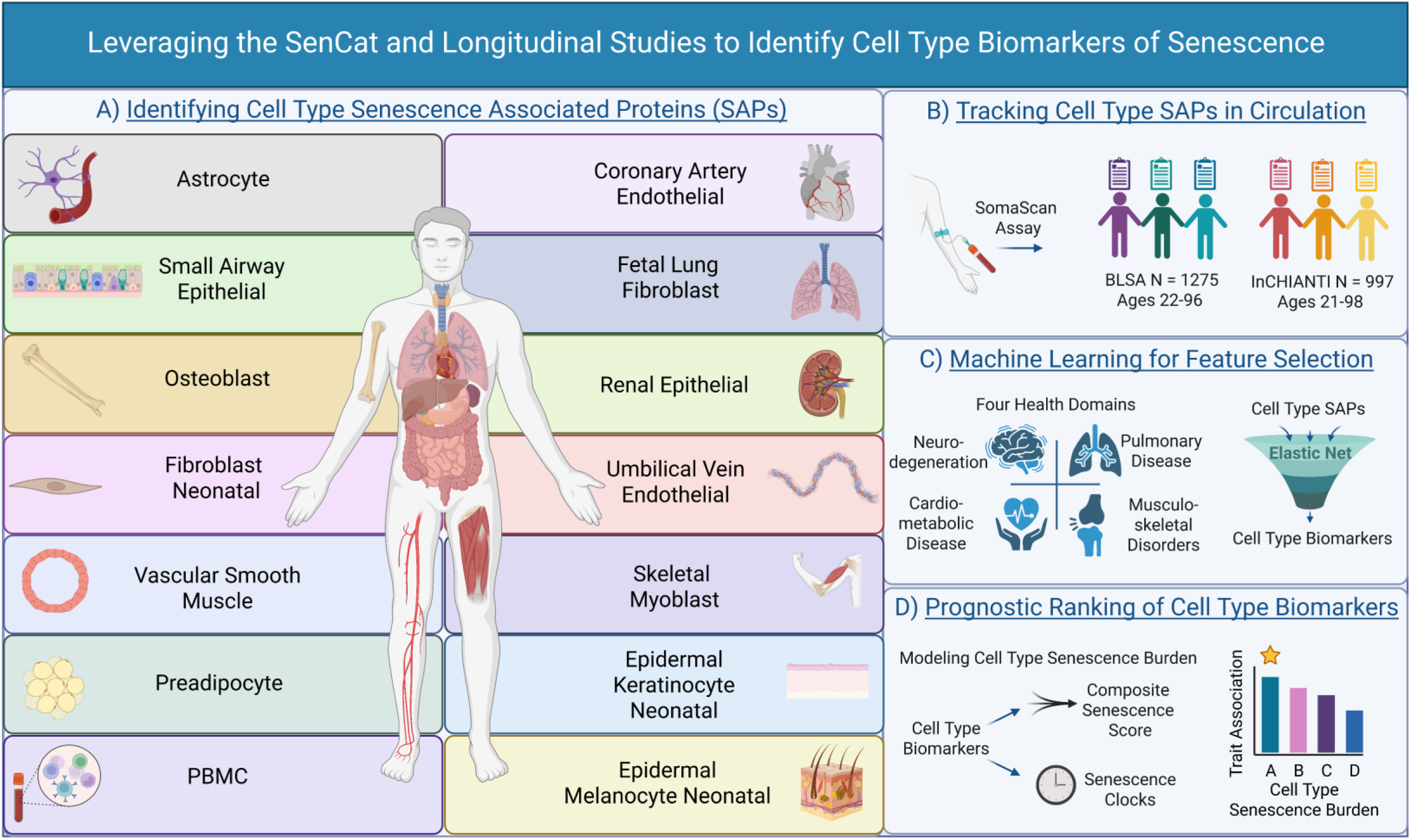
Leveraging the SenCat and Longitudinal Studies to Identify Cell Type Biomarkers of Senescence. Study design for development of core and cell type-specific senescence signatures and associations with health status: A) Senescence signatures from 14 cell types reported in the Senescence Catalog (SenCat) were used to identify both core and cell type-specific proteomic senescence signatures. B) The circulating plasma proteomes of the BLSA and InCHIANTI studies were quantified and senescence signatures were tracked. C) Machine learning was used to identify circulating cell type senescence biomarkers that predict a diverse set of aging-associated phenotypes corresponding to multiple health domains. 4) The senescence burden was modeled using cell type senescence biomarkers and ranked by trait association and prediction potential.

**Figure 2:**
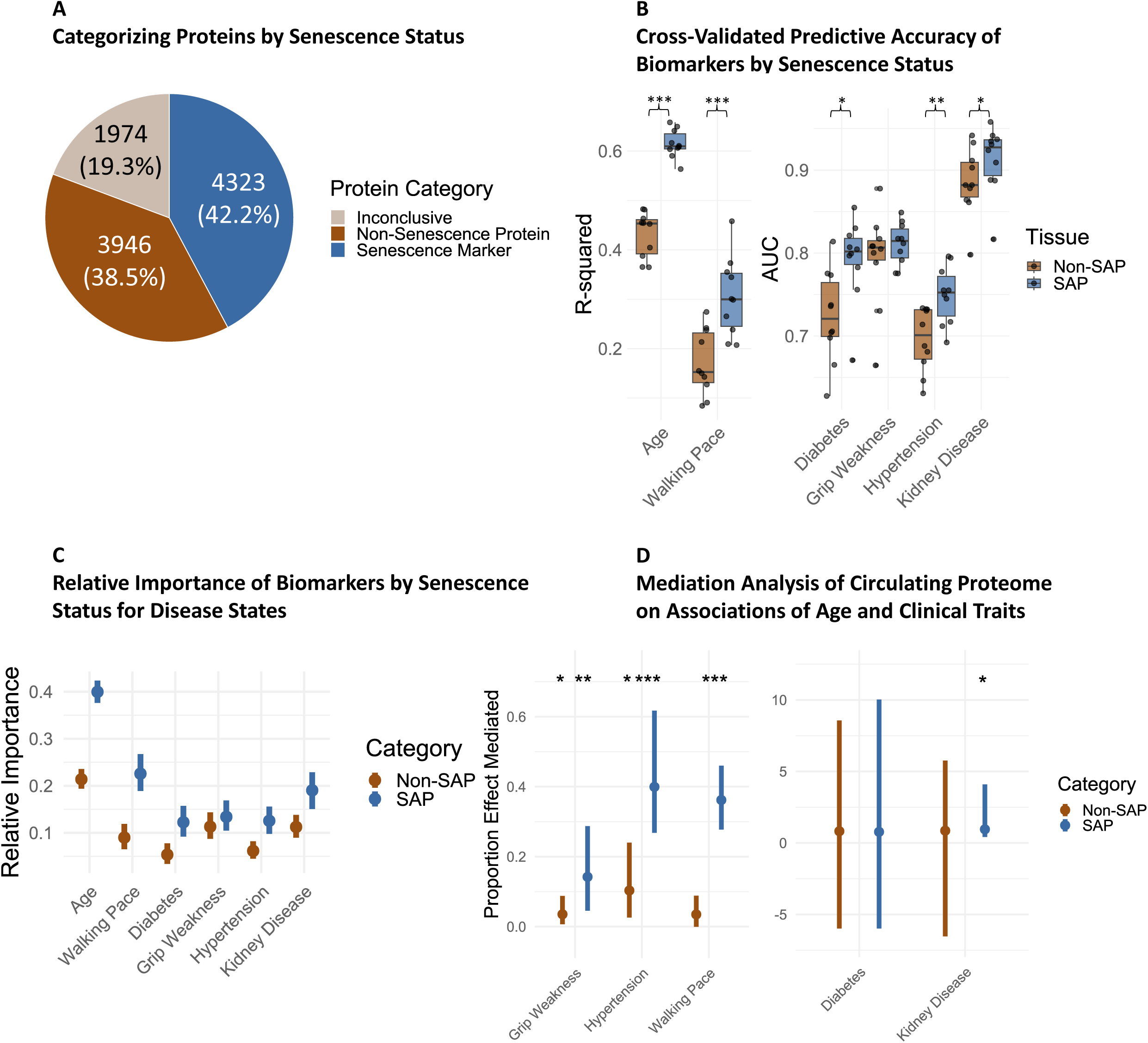
Senescence Proteins Outperform Non-Senescence Proteins as Biomarkers. A) Senescence Catalogue proteins by senescence status. B) Cross-validated R-Squared values were identified across ten iterations (90% train, 10% test) for clinical parameters. To normalize different pool sizes, only the top 25 ENSPs were used for each group of proteins, which were condensed into a mean composite score. Paired T-Tests were used to compare SAP vs. Non-SAP parameter prediction, which showed significant differences among most traits. C) Relative importance analysis of composite SAP or Non-SAP proteins revealed higher relative importance of the SAP score. D) Mediation analysis was conducted to determine if either SAP or Non-SAP composite scores mediate the relationship between age and several clinical traits while controlling for sex, race, and the alternative composite score. The proportion of the effect of age on the clinical traits mediated by each composite score is shown, with p-values on top of each. Error bars denote a 95% confidence interval.

### Senescence markers are detected in the BLSA and Inchianti

Senescence and non-senescence proteins were examined for clinical associations in circulating plasma in a cohort of 1275 in the BLSA (ages 22-96, mean 67.52) and 997 participants in the InCHIANTI study (ages 21-98, mean 66.31). A single proteomics sample from each participant was examined in this study. Proteins were quantified in both cohorts using the SomaScan assay (Somalogic Inc., Boulder, CO) which identifies proteins using a Slow Off-Rate Modified Aptamer system. The 7k SomaScan was used to quantify approximately 7000 proteins in the BLSA, and the 1.3k SomaScan assay was used to quantify approximately 1300 proteins in InCHIANTI. Clinical characteristics are available for the BLSA cohort in **Supp. Table 2**, and the InCHIANTI cohort in **Supp. Table 3**. Of the proteins identified in the SenCat, 3944 were also detected in the BLSA and 712 were detected in InCHIANTI. The percentages of total SAPs, Non-SAPs, and inconclusive proteins identified in the BLSA were 49%, 32%, and 19% respectively. The corresponding percentages in InCHIANTI were concordant at 50%, 32%, and 18% respectively. A Fisher’s Exact test revealed that there was not a meaningful difference between these quantifications between studies (**Supp. Table 4**), highlighting their suitability for cross-study validation. A protein index listing proteins detected in both the SenCat and in the plasma of participants of each longitudinal study, including their senescence status is included for the BLSA (**Supp. Table 5**) and InCHIANTI (**Supp. Table 6**).

### Senescence-associated proteins outperform non-senescence proteins in trait prediction

To compare the broad clinical relevance of SAPs versus all other circulating proteins, SAPs and non-SAPs were used for modeling to predict clinical parameters in the BLSA cohort including age, walking pace, diabetes status, and hypertension, among others. To prevent overfitting, elastic net modeling was used to perform feature selection from each protein group. Elastic Net Selected Proteins (ENSPs) were identified from each group and for each clinical trait while controlling for age, sex, and race as covariates. To account for different protein pool sizes, only up to the top 25 proteins by effect size were selected from each group, and each group of selected proteins was condensed into a mean composite score of either SAPs or Non-SAPs to represent the senescence or non-senescence burden. Ten rounds of cross-validation were then performed (90% Train, 10% Test) using linear or logistic regression models that were trained using only the trait-specific composite score per group to predict each clinical parameter. Notably, trait prediction performed better with the senescence composite score than the non-senescence score for most traits including age, walking pace, diabetes, kidney disease, and hypertension (**Fig. 2b**). The distributions of the differences between the paired SAP and Non-SAP trait prediction were determined to be approximately normal using Shapiro Wilk tests (p-value > 0.05), and were therefore suitable for comparison using a paired t-test, which indicated significant or nearly significant differences between the SAP and Non-SAP prediction for most traits (**Supp. Table 7**). When comparing clinical relevance of SAPs and non-SAPs in either younger (<65) or older (65+) groups, SAP again generally outperformed the non-SAP, particularly in the older group (**Supp. Fig. 1b-c**). Similar differences in predictive accuracy were found in the InCHIANTI study (**Supp. Fig. S1A, S1D, S1E**). Additionally, when SAP and non-SAP composite scores were combined into one model, the SAP scores obtained higher relative importance for these traits (**Fig. 2c**). Last, the SAP composite score better mediated the relationship between age and these traits than the non-SAP score, while controlling for sex, race, and the alternative composite score (**Fig. 2d**). While these results do not necessarily establish a causal role that senescence plays in aging trajectories, they suggest that senescence-associated protein signatures may outperform non-senescence-associated proteins as clinically relevant biomarkers.

Additionally, the SAP and Non-SAP groups were assessed for their ability to predict traits in one study after training on data from the other study. Elastic net was performed on both studies, and only those features selected in both studies were combined into a single model. From here these models were either trained on the BLSA and then used to predict traits in InCHIANTI, or trained on InCHIANTI and used to predict traits in the BLSA. The SAP models generally had higher predictive accuracy than the Non-SAP models, and models trained on the BLSA were often slightly more accurate at predicting traits in InCHIANTI than models trained on InCHIANTI to predict traits in the BLSA. This again suggests that the circulating SAP signature may be slightly more clinically informative than the circulating Non-SAP signature. Selected features in both studies along with their performance metrics are tabulating in **Supp. Table 8**.

### Sex-specific differences and temporal patterns of senescence throughout the lifespan

Previous studies have found that the plasma proteome undergoes nonlinear changes that become accelerated at various decades of life in human cohorts.^35^ Following this line of inquiry and using Differential Expression Sliding Window Analysis (DE-SWAN), a tool developed by Lehallier et al,^36^ SAPs were similarly investigated in the BLSA to determine if they also undergo nonlinear changes. To address imbalances in the BLSA cohort age distribution, only those in the age range of 50-90 were used in the analysis, and the cohort was downsampled to include the same number of randomly selected individuals by decade of life (N=120 per decade, 480 total, 50% male/female). A seed of 1 was used with the slice_sample function of the dplyr package for downsampling. Standardized differences of age clinical traits between the selected group and the eligible group (ages 50-90) were compared, and SMD comparing the two groups was generally low (abs(SMD) <0.12) except for age (SMD = -.321), which indicates that the random sampling by decade indeed corrected an uneven age distribution (**Supp. Table 9**). The age distributions of the entire BLSA cohort (n=1275, ages 22-96), the eligible cohort (N = 1093, ages 50-90), and the selected group (N=480, ages 50-90) are shown in **Supp Fig. 2a**. Differentially abundant SAPs were identified at 1-year intervals from 60-80 by comparing 10-year age windows. For example, differentially expressed proteins at 60 were identified by comparing those aged 50-60 to those aged 60-70, and this was repeated for each 1-year interval from 60 to 80. Of the ages surveyed, the highest number of both SAPs and Non-SAPs were elevated at 78, corroborating previous findings showing peak upregulation at this exact year,^35^ and suggesting that broad proteomic changes occur in this decade of life. When SAPs were analyzed separately by sex, an interesting pattern emerged: though proteomic changes appeared to peak earlier, at age 72, in females compared with age 78 in males, the number of upregulated SAPs at this peak was much greater in males (N = 77) compared with females (N = 45) (**Fig. 3a**). Further, ontology analysis of all senescence proteins increased each year revealed pathways related to immune cell proliferation and sphingolipid metabolism in the early 60s, and these upregulated processes shifted to cytokine production and protein processing by the late 70s (**Supp. Fig. 2b**). When overrepresentation analysis was conducted on collectively elevated SAPs in ages 60-80 in males and females separately, the two groups again diverged; in males, processes related to immune cell activation were overrepresented, while processes related to extracellular matrix organization were overrepresented in females (**Supp. Fig. 2c**). These insights highlight the shifting nature of the senescence burden in later life and its potential asymmetry in emergence between males and females.

**Figure 3:**
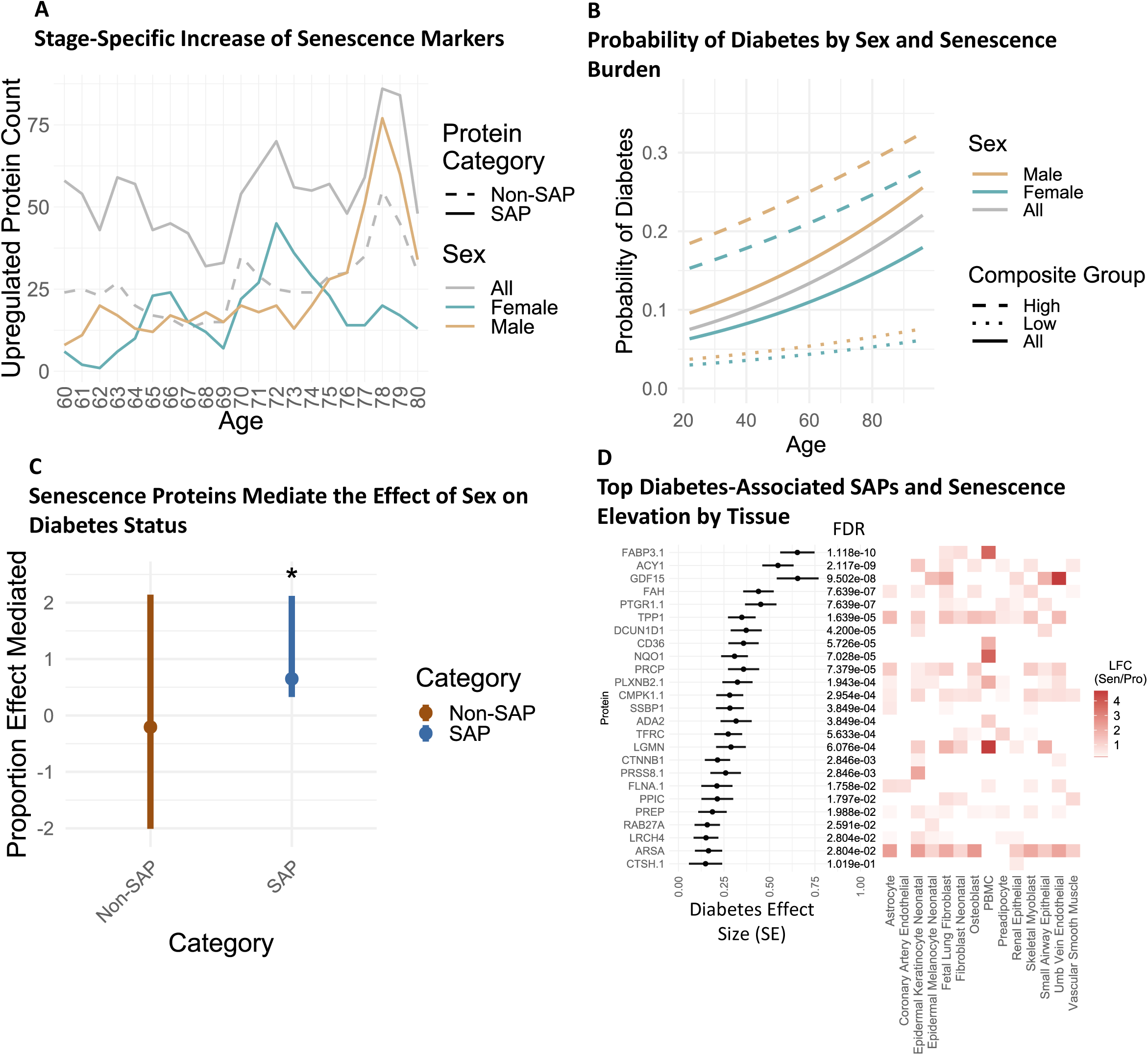
Senescence Markers Show Non-Linear Changes Throughout the Lifespan and Mediate the Relationship Between Sex and Disease Status. A) SAPs elevated by year in the BLSA using the Differential Expression Sliding Window Analysis (DE-SWAN) method comparing 10-Year windows per year from 60 to 80. B) The probability of diabetes by sex and highest or lowest quartile composite senescence burden using logistic regression with models: diabetes ∼ age, diabetes ∼ age + sex, or diabetes ∼ age + sex + composite SAPs score. C) The proportion effect mediated of the relationship between sex and diabetes status using either SAPs or Non-SAPs selected via elastic net modeling and condensed into a composite score. Error bars denote a 95% confidence interval. D) Logistic regression effect size of SAP ENSPs for diabetes using the formula: diabetes ∼ protein + age + sex + race, and their respective log fold-change (LFC, Sen/Pro) by cell type. *For Sen/Pro LFC, only proteins significantly elevated in each cell type using two induction methods are depicted by their average LFC across both induction methods.

On average, men and women exhibit differences in mortality^37^ and the onset and severity of diseases such as diabetes.^38^ Additionally, men and women may differ in their senescence susceptibility and senescence burden.^39^ In the BLSA cohort, logistic regression revealed that diabetes increased in likelihood with age, and exhibited sex-specific differences in prevalence, with males being more likely to have the disease than females (**Fig. 3b**). The role that senescence markers play in the association of sex with diabetes status was investigated using mediation analysis. Elastic Net modeling was first used to identify SAPs and Non-SAPs associated with diabetes status, which were limited to 25 features from each group and condensed into a mean composite score per participant in the BLSA. Mediation analysis revealed that the SAP composite score mediates the association between sex and diabetes status in the BLSA, while the non-SAP score does not (**Fig. 3c**). SAP ENSPs significantly associated with diabetes include FABP3, ACY1, GDF15, FAH, and PTGR1 among others (**Fig. 3d**). Fatty Acid Binding Protein 3 (FABP3) was most strongly associated, which corroborates previous findings that FABP3 plays a role in diabetes-associated lipotoxicity of pancreatic beta cells^40^ and is associated with diabetes when quantified in circulation in humans.^41^ Additionally, when senescence-associated ENSPs for diabetes status were condensed into a composite score, those in the highest and lowest quartile composite scores diverged more strongly in lifetime susceptibility to diabetes than was seen between males and females generally (**Fig. 3b**). These findings suggest that circulating SAPs not only accumulate at varying rates by sex across the lifespan but importantly may also play a causal role in the associations between sex and disease trajectories.

### Core senescence proteins predict clinical traits in BLSA and InCHIANTI

Many cell type senescence signatures broadly overlapped, and any given SAP was upregulated in as few as one and as many as all fourteen senescent cell types (at least two induction methods for each cell type). Despite broad heterogeneity in senescence proteins by cell type and induction method, a group of SAPs was identified as core senescence markers in multiple cell types. This core consisted of 315 proteins that were identified as senescence markers in at least 9 of the 14 cell types and were investigated further for clinical associations. The top 25 core proteins implicated in age were identified using linear modeling while controlling for sex and race and shown by their association with age and their average log2 fold-change (senescence versus proliferating) by cell type, including GM2A, CTSB, LTA4H, and PLCD1 with the highest age association, among others (**Fig. 4a**). Up to 25 core proteins implicated in several clinical parameters were selected via elastic net, which collectively had moderate to high cross-validated predictive ability in the BLSA and InCHIANTI studies for some clinical parameters including age, kidney disease, and diabetes (**Fig. 4b**).

**Figure 4:**
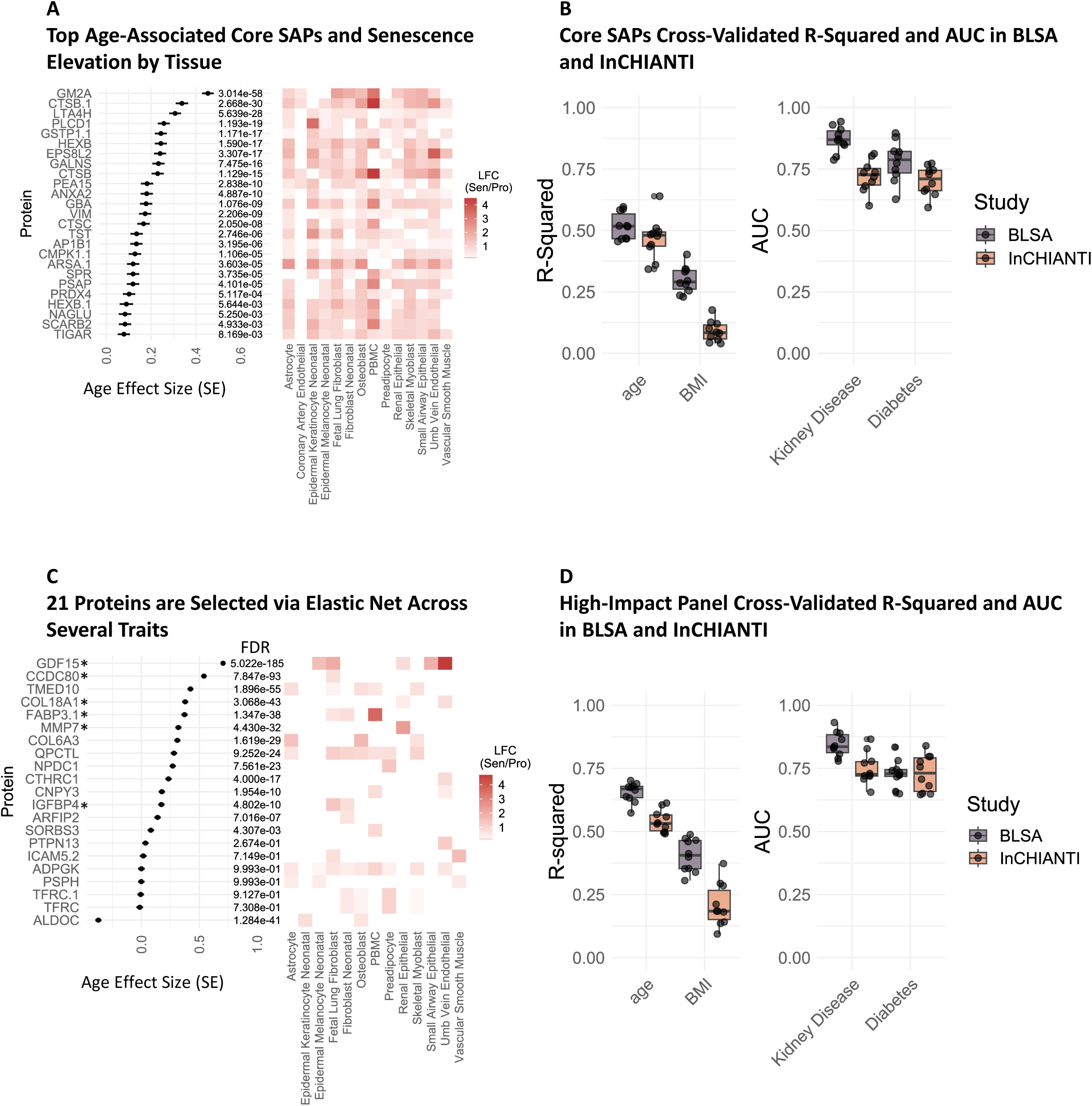
Core SAPs Predict Clinical Parameters. A) 315 proteins were elevated in senescence following two induction methods in at least 9 of the 14 SenCat cell types and were identified as core SAPs. A) The top 25 age-associated core SAPs using linear modeling with the formula age ∼ protein + sex + race, and their respective average Log2 Fold Change (LFC, Sen/Pro) between two induction methods for each cell type. B) Elastic net modeling was used to select up to 25 core SAPs implicated in several clinical parameters, and their collective cross-validated predictive potential across 10 iterations in the BLSA and InCHIANTI studies was identified using linear or logistic regression with the formula: parameter ∼ protein 1 abundance + protein 2 abundance … + protein 25 abundance. C) Elastic net selected proteins were identified for ten clinical parameters and labeled as a High Impact Panel (HIP). *Indicates detection in the InCHIANTI study. The association with age for the HIP in the BLSA using linear modeling with the equation age ∼ protein + sex + race, and the LFC (Sen/Pro) of the HIP was found by cell type in the SenCat. D) The cross-validated predictive potential across 10 iterations for a trait panel was determined for the HIP in the BLSA and InCHIANTI studies using linear or logistic regression and the formula: parameter ∼ protein 1 abundance + protein 2 abundance … + protein 25 abundance. *For Sen/Pro LFC, only proteins significantly elevated in each cell type using two induction methods are shown by their average LFC across both induction methods.

Additionally, senescence markers derived from any of the 14 cell types were compiled and examined for clinical associations in the BLSA. Elastic net modeling was used to identify SAPs implicated in a panel of clinical parameters that span a broad range of health domains, including age, BMI, appendicular lean mass, spinal bone density, maximum grip strength, walking pace, lung function, kidney function (eGFR), cognition score, and a 44-component frailty index. Proteins selected in at least half of the traits were identified as a High-Impact Panel (HIP), which are listed in the BLSA (**Supp. Table 5**) and InCHIANTI (**Supp. Table 6**). The HIP components showed varying associations with age in the BLSA, with GDF15 the most strongly associated, and were senescence markers in the SenCat across a variety of cell types (**Fig. 4c**). Cross validation revealed that these proteins collectively predict several clinical traits, including age, BMI, kidney disease, and diabetes in both the BLSA and InCHIANTI (**Fig. 4d**). An important consideration when comparing predictive capability of the SAP in each cohort is that far fewer proteins were measured in InCHIANTI, and thus fewer SAP are measured. In many cases the models remain robustly significant, albeit lower performing than in the BLSA. Collectively, this suggests the clinical relevance of the core SAPs that, along with the HIP, could potentially serve as biomarkers in future clinical settings.

### Cell type-exclusive senescence markers map to their corresponding health domain

Senescence is known to accumulate across all tissues with age. Notably, recent studies have demonstrated that organ-specific plasma proteomic signatures, generated based on tissue-specific proteins, predict the biological aging of organs and associate with organ-specific disease risks in humans.^34^ To similarly investigate senescence-driven age associations by tissue, senescence signatures were identified for each cell type (**Supp. Fig. 3a**). For this and subsequent analyses, neonatal cell types were excluded due to their unlikely contribution to health domain-specific aging signatures in adult humans, and the remaining 9 cell types present in adults were included in the analysis; additionally, a core group of SAPs (N = 300) specific to these 9 cell types were identified as elevated in at least 6 of the 9 cell types. To further ensure unique signatures for each cell type, exclusive senescence signatures were identified by cell type using two criteria: 1) statistically significant (positive LFC Sen/Pro, adjusted p-value < 0.05) increased abundance in two induction methods for a given cell type and 2) not a senescence marker in any other cell types within SenCat. Importantly, the cell type-exclusive label for each signature indicates its exclusiveness in the SenCat, and does not indicate that this signature is sourced exclusively from the given cell type when quantified in circulation. Though the SenCat provides evidence that these cell types contribute to their respective signatures in circulation, it is likely that these signatures originate from multiple sources throughout the body. For each group of SenCat cell type-exclusive SAPs, linear modeling was used to identify their association with age while controlling for sex and race. Interestingly, of the selected cell types, osteoblasts contained the largest pool of SAPs (1370 proteins), while coronary artery endothelial cells contained the smallest (120 proteins). Proportions of age-related SAP were identified by cell type and revealed that among SAPs unique to each cell type, coronary artery endothelial cells contained the highest proportion of positively age-related senescence proteins, while skeletal myoblasts contained the lowest proportion (**Supp. Fig. 3b**). Of all SAPs, GDF15, TAGLN, CCDC80, and PDLIM3 were the most strongly associated with age, among others, and were senescence signatures across multiple cell types (**Supp. Fig. 3c**).

To further examine whether the cell type-exclusive senescence signatures derived from the SenCat are present in their corresponding healthy cell types in human tissues, these signatures were examined in an external dataset, the Human Protein Atlas (HPA). The HPA examines protein expression using immunohistochemistry in 45 human tissues, with some being examined across multiple sections and representing multiple cell types. Tissues for cross comparison were selected by finding those that roughly match to the same organ system of cell types represented in the SenCat, while limiting the analysis to those tissues with multiple cell types examined. Ultimately, cell type-exclusive senescence signatures of small airway epithelial, renal epithelial, PBMCs, and astrocytes were examined in matching tissues, including the lung, bronchus, kidney, lymph node, bone marrow, and cerebellum. Protein expression in the HPA is reported as not detected, low, medium, or high, and for each cell type-exclusive SAP signature the proportion of proteins from that signature that were reported as medium or high was found. Strikingly, among all tissues, the three sections with the highest proportion of medium or highly expressed proteins from the small airway epithelial signature were found in the bronchus and lung (**Supp. Fig. 4a**). Similar patterns were found for the other three cell type SAPs, such as the section with highest proportion of renal epithelial medium or high expression was in the kidney, with the second and third being in the bronchus and lung. The section with highest proportion of PBMCs SAP expression was also in the lung, though notably in the section represented by macrophages. Last, the astrocyte SAPs showed the highest proportion of medium/high expression in the cerebellum. Collectively, this analysis of the Human Protein Atlas confirms, for multiple cell types, the presence of a majority of SenCat’s cell-type-exclusive protein signatures at medium or high levels in their corresponding healthy cells and tissues. The strength of the SenCat database is the further identification of cell-type exclusive signatures in the senescent state, enabling the identification of plausible candidate cell type-specific senescence signatures in circulation.”

Each set of SenCat cell type-exclusive SAPs was investigated for its clinical relevance across a variety of traits. For each trait, elastic net models were used to identify up to 25 cell type-exclusive SAPs from each cell type implicated in that trait, which were each condensed into a mean composite score to represent each cell type senescence burden. These composite senescence burden scores were then examined for clinical associations in the BLSA and InCHIANTI using linear or logistic regression models, and any unique clinical relevance by cell type was identified. Clinical parameters and disease statuses were collected to represent health status across a broad range of health domains: this includes body composition and diabetes status to represent cardiometabolic health; MMSE test cognition score and dementia status to represent neurodegeneration; appendicular lean mass, grip strength, and walking pace to represent musculoskeletal health; and forced expiratory volume and COPD status to represent pulmonary health, among others. An unbiased view of the cell type composite score associations with clinical traits and disease status is shown for BLSA and InCHIANTI (**Supp. Fig. 5a-d)**.

In many instances, cell type senescence signatures were robustly associated with traits across studies. Strikingly, PBMC and renal epithelial senescence burden most strongly associated with a multi-component frailty index in the BLSA (**Fig. 5a**) and were both in the top three, along with astrocytes, in InCHIANTI (**Supp. Fig. 5c**). In the BLSA, among the PBMC SAPs selected via elastic net to represent senescence burden, FABP3, CNYP3, HAVCR2, GPNMB, and LGALS9 were the most strongly associated with the frailty index (**Supp. Fig. 6a**). Senescence in circulating T-cells, a component of PBMCs, has also been implicated in diabetes in human cohorts.^42^ Corroborating this, PBMC senescence burden had the most significant association with diabetes status in both the BLSA (**Fig. 5b**) and InCHIANTI (**Supp. Fig. 5d**). Among the PBMC elastic net selected senescence signatures for diabetes, FABP3, CPM1, CD36, F9, and ADA2 had the strongest association with diabetes in the BLSA (**Supp. Fig. 6b**). Diabetes was a component of the frailty score, and so a sensitivity analysis was conducted to determine if diabetes contributed to the association between PBMC senescence score and frailty. A frailty score without diabetes was subsequently constructed, and cell type SAP were similarly modeled, and again, the PBMC cell type senescence burden was the most strongly associated (**Supp. Fig. 6c**). This suggests that the PBMC senescence burden is associated with frailty irrespective of the contribution of diabetes status.

**Figure 5:**
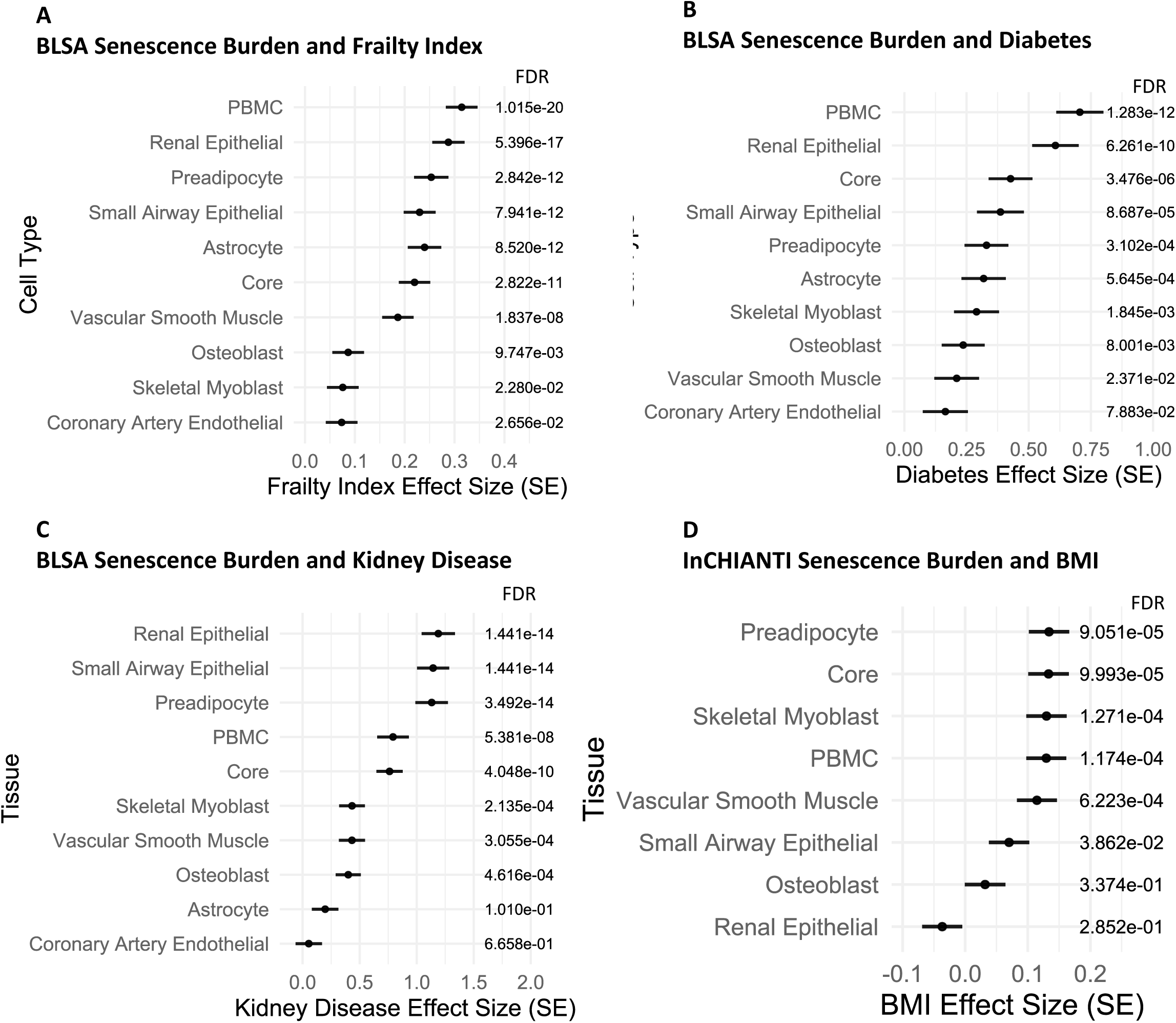
The Immune Senescence Signature is the Strongest Cross-Study Correlate with Frailty and Diabetes. Cell Type Senescence Burdens Associate with Clinical Parameters in the Corresponding Health Domain. Up to 25 elastic net selected SAPs were selected from cell type-exclusive senescence signatures within SenCat as implicated in a frailty index or separately for diabetes status, which were each compiled into a mean composite score to represent cell type senescence burden. A) The association between the cell type senescence burden and frailty index was found with Pearson linear modeling using the formula: frailty index ∼ Composite Score + age + sex + race. B) The association between the cell type senescence burden and diabetes status with logistic regression using the formula diabetes ∼ Composite Score + age + sex + race. C) The association between the cell type senescence burden and kidney disease status in the BLSA using logistic regression with the formula: kidney disease status ∼ composite score + age + sex + race. D) The association between the cell type senescence burden and BMI in InCHIANTI using linear modeling with the formula: BMI ∼ composite score + age + sex.

Notably, SenCat cell type-exclusive SAPs were often best correlated with clinical parameters that map to their respective health domain. For instance, the renal epithelial senescence signature was the most strongly associated with kidney disease in the BLSA (**Fig. 5c**), with COL18A1, PXDN, MMP7, DCLK1, and SERPINB8 among the renal epithelial SAPs selected via elastic net most strongly associated with kidney disease using logistic regression (**Supp. Fig. 6d**). In InCHIANTI, senescence burden of preadipocytes, cells critical for adipose tissue function and lipid regulation, were the most highly associated with BMI (**Fig. 5d**). This result corroborates previous findings that senescence markers in adipose tissue positively correlate with BMI in humans and show concomitant loss of stem cell potential,^43^ an effect thought to contribute to obesity-related morbidities,^44^ and suggests that these adipose-related markers may also be detectable in circulation. Among preadipocyte SAPs selected via elastic net, DCN, LDLR, and GOT1, among others, best correlated with BMI in InCHIANTI (**Supp. Fig. 6e**). Interestingly, decorin (DCN), which plays a role in the extracellular matrix, was previously found to be highly expressed in adipose tissue of a preclinical model, and associated with waist to hip ratio when quantified in circulation in an independent human cohort.^45^ Additionally, senescence burden composite scores were compared for predictive ability of clinical parameters and again often had differential predictive ability that mapped to the corresponding health domain. Small airway epithelial senescence burden, for instance, had the highest cross-validated prediction ability for COPD status in the BLSA, with the ENSPs HLA-G and NAB1 being the most strongly associated with COPD (**Supp. Fig. 7a,b**). Additionally, skeletal myoblast senescence burden best predicted max grip strength in InCHIANTI, with PLAUR among the ENSPs showing a significant inverse association (**Supp. Fig. 7c,d**).

### Senescence signatures are associated with longitudinal outcomes

Senescence signatures, when quantified in circulating plasma, have previously been associated with longitudinal outcomes such as mortality.^11^ A subset of senescence signatures from any cell type implicated in mortality in the BLSA were identified via elastic net modeling, and Cox proportional hazards modeling revealed SAPs significantly associated with mortality. PLTP, MMP7, and ENGASE, among others, were the most significantly associated with mortality, and are expressed as a senescence marker in several cell types (**Fig. 6a**). The top ranking protein PLTP, important for lipid processing and implicated in atherosclerosis, has been previously associated with lower survival rates in humans when quantified in the plasma.^46^ Notably, when examined using Shoenfeld residuals, most of the SAPs satisfied the proportional hazards assumption necessary for cox proportional hazards modeling, while some conversely showed a time-dependent effect on mortality (**Supp. Table 10**). When condensed into a mean composite senescence burden score, those in the BLSA with expression of these ENSPs in the upper quartile showed higher lifetime mortality than those in the lowest quartile, particularly in later life (**Fig. 6b**). A steep divergence in mortality probability was observed between the two groups beginning at age 50, but notably the smaller sample size at earlier ages limits inferences that can be made from this. Cell type composite senescence burden scores were similarly identified using up to 25 ENSPs unique to each cell type and compared for longitudinal clinical associations. Unlike the single mortality-associated SAPs, none of the cell type composite scores violated the proportional hazards assumption (**Supp. Table 10**), indicating that the composite scores may be more suitable for predicting longitudinal outcomes. When the cell type composite scores were compared, PBMC senescence burden had the highest degree of association with diabetes onset (**Fig. 6c**). When PBMC ENSPs were condensed into a mean composite senescence burden score, those in the BLSA with the upper quartile expression displayed a higher lifetime likelihood of developing diabetes than those in the lower quartile (**Fig. 6d**). PBMC senescence burden was notably also the most strongly associated with kidney disease onset (**Supp. Fig. 8a**), and hypertension onset (**Supp. Fig. 8b**). This suggests that not only can the circulating senescence burden, particularly when derived from immune cells, shed light on present disease status but can also indicate health trajectories throughout the lifespan. Elastic net selected proteins were compiled into a single table showing features selected by cell type SAP, trait, and study (**Supp. Table 11**).

**Figure 6:**
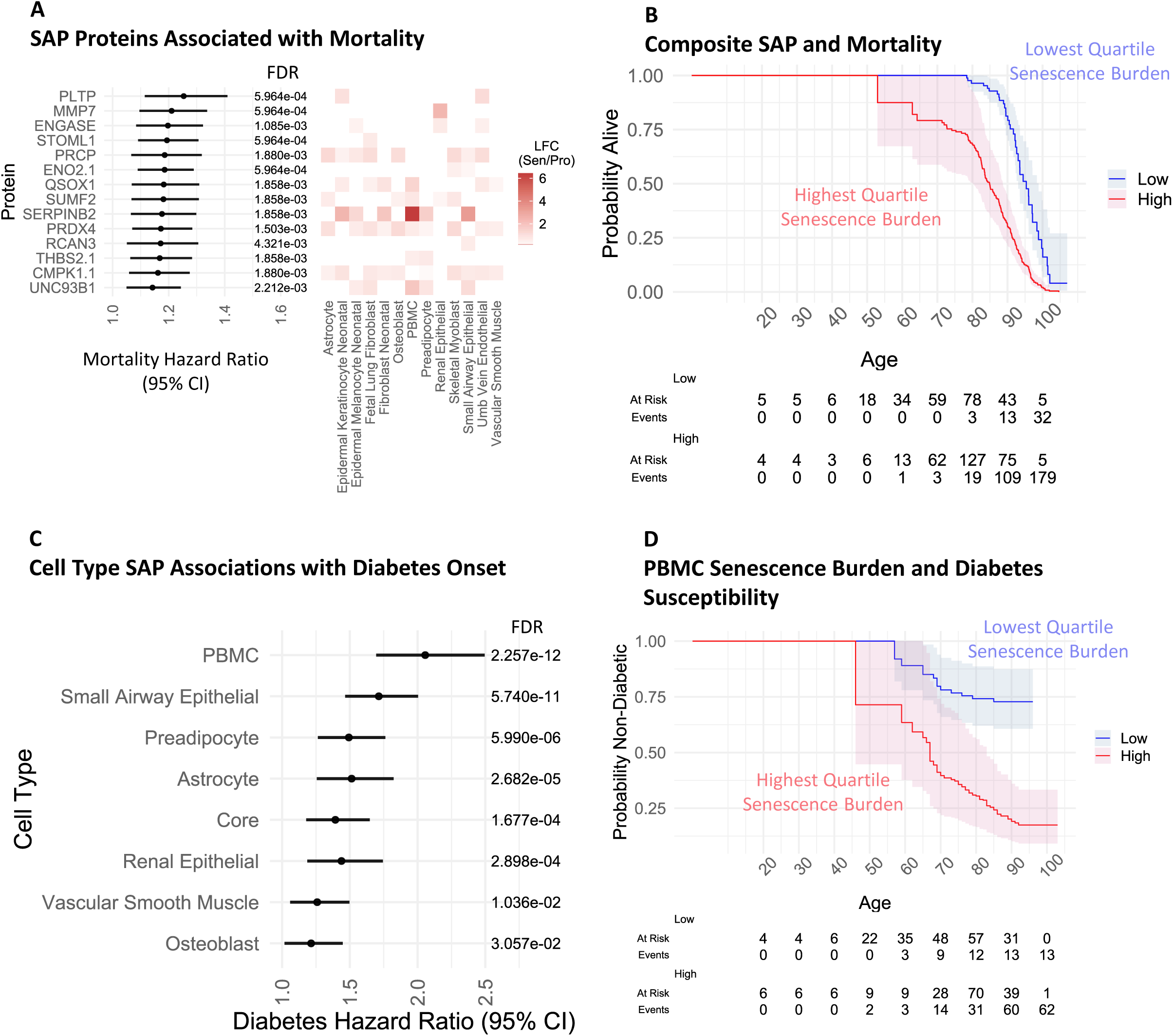
Longitudinal Clinical Associations of Senescence Signatures. A) Among all SAPs identified in at least two induction methods for each cell type in the SenCat, elastic net modeling revealed senescence signatures associated with mortality. Linear modeling revealed each signature association with age using the formula age ∼ protein + sex + race, and their average log2 fold-change (LFC Sen/Pro) in each cell type across two induction methods is shown. B) Tracking longitudinal health trajectories reveals higher mortality among those in the highest quartile mean composite senescence score of these SAPs compared to those in the lowest quartile. C) Up to 25 elastic net selected SAPs were selected from cell type senescence signatures as implicated in a diabetes onset, which were each compiled into a mean composite score to represent cell type senescence burden. The association between the cell type senescence burden and diabetes onset via cox proportional hazards modeling in the BLSA using the formula: event/age or last age ∼ composite score + sex + race. D) Tracking longitudinal health trajectories reveals higher diabetes probabilities among those in the highest quartile composite score of PBMC SAPs compared to those in the lowest quartile. *For Sen/Pro LFC, only proteins significantly elevated in each cell type using two induction methods are depicted by their average LFC across both induction methods.

### Deriving cell type “seno-ages” from senescence-associated proteins

Recent work demonstrated that plasma proteins can model organ aging and “age gaps”, a measure of an individual’s biological age relative to a cohort average, and organ-specific age gaps were predictive of organ aging phenotypes.^34^ To expand these efforts to the context of senescence-associated aging, we employed a similar approach to model organ ages by using SAP to calculate the “seno-age” of each cell type. Cell type seno-ages were calculated using SenCat cell type-exclusive SAPs selected via elastic net, which were used within a linear model to predict the cell type seno-age for each individual. Cell type seno-ages showed varying trajectories throughout the lifespan. Small-airway epithelial and the core, among others, had the strongest relation with chronological age in both BLSA and InCHIANTI, particularly between 50 to 80 years of age (**Supp. Fig. 9a,b**). Some seno-ages were moderately intercorrelated throughout the lifespan, in particular the core (correlation coefficients ranging from r = 0.2 – 0.7 with all cell types), PBMC (r = 0.15 – 0.7), and astrocyte (r = 0.16 – 0.72) seno-ages in both BLSA and InCHIANTI (**Supp. Fig. 9c,d**). Expanding from this, cell type seno-age gaps were calculated as the residual between the trend line of each individual’s chronological age and their predicted seno-age, which theoretically identifies individuals who appear by their circulating proteome to be older (thus a positive seno-age gap) or younger (thus a negative seno-age gap) than their chronological age (**Supp. Fig. 10a**). These cell type seno-age gaps were then investigated for clinical associations in the BLSA and InCHIANTI (e.g. kidney disease, **Supp. Fig. 10b**).

### Cell type seno-age gaps uniquely associate with organ aging phenotypes

Cell type seno-age gaps were next investigated for associations with clinical traits and disease using linear or logistic regression modeling in the BLSA and InCHIANTI. Seno-age gaps showed broad and statistically significant trait and disease state associations in both the BLSA (**Supp. Fig. 11a,b**) and InCHIANTI (**Supp. Fig. 11c,d**). Notably, some cel type seno-age gaps were associated with their respective health domain. For example, skeletal myoblast seno-age gap was ranked as the most significant inverse association with appendicular lean mass in the BLSA (**Fig. 7a**). This corroborates previous findings that link senescence-related inflammation with loss of muscle stem cell regenerative function,^47,48^ an effect mediated by senolytic treatment with rapamycin or spermidine.^49^ Interestingly, most of the skeletal myoblast SAPs that were selected to model cell type seno-age gap were inversely associated with age in the BLSA, including OTULIN, RAC2, ASL, and CLINT1, among others (**Fig. 7b**). Among previously reported organ aging signatures in plasma, brain age-gap was highlighted as a predictor of dementia severity and Alzheimer’s Disease.^34^ Consistent with previous findings, our analysis revealed that astrocyte seno-age gap, the only brain cell type in the SenCat, most strongly associated with dementia status in InCHIANTI (**Fig. 7c**). The contributing proteins to this association include B2M, PSMA2, CFL1, PSMD7, and NOTCH2 (**Fig. 7d**). This corroborates previous studies that found that plasma levels of B2M are associated with worse cognitive performance and Alzheimer’s Disease,^50^ and additionally establishes novel biomarker candidates for neurodegeneration.

**Figure 7:**
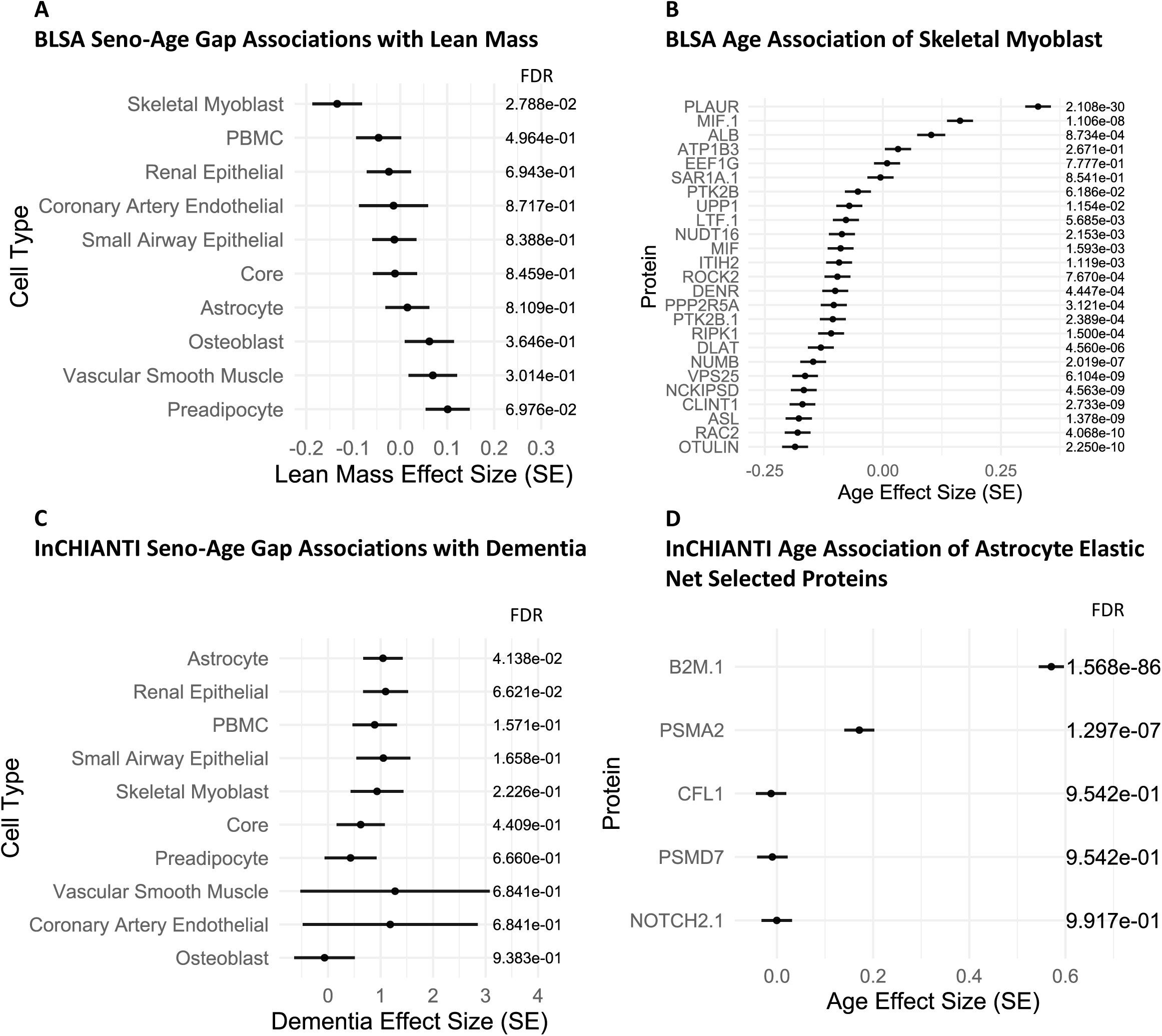
Senescence Age-Gap Associations with Clinical Parameters. Seno-Age Gap was derived using up to 25 Elastic Net Selected Proteins to model age from each cell type-exclusive set of SAPs within SenCat. and subsequently comparing this modeled age with chronological age. A) The association between the cell type Seno-Age Gap and appendicular lean mass in the BLSA using linear modeling with the formula: lean mass ∼ composite score + sex + race. B) Age associations via linear modeling for Skeletal Myoblast SAPs selected via elastic net modeling in the BLSA using the formula: age ∼ protein + sex + race. C) Seno-Age Gap associations were similarly calculated for InCHIANTI, and cell type Seno-Age Gap associations with Dementia were calculated using logistic regression using the formula: dementia status ∼ composite score + sex. D) Dementia associations via logistic regression for Astrocyte SAPs selected via elastic net modeling in InCHIANTI using the formula: dementia status ∼ protein + sex.

## DISCUSSION

This study undertook one of the most comprehensive investigations into senescence markers in circulating plasma to date, and the first investigation of cell type-specific (senotype-specific) senescence signatures and their possible clinical relevance across two human cohorts. Senescence proteins were associated with diverse aging phenotypes in the BLSA and InCHIANTI, and outperformed other (non-senescence-associated) circulating proteins in predicting many clinical parameters, including age, walking pace, and hypertension. Further, senescence proteins better mediated the relationship between age and these clinical parameters, suggesting plasma senescence burden as a potential driver of age-associated trait changes.

Our study adopts a prior framework that utilized plasma protein signatures to model organ health and biological aging.^34^ Expanding on this, we present the first resource of senotype-enriched plasma protein profiles of senescence based on 14 different cell types, proposing both core and cell type protein signatures for prediction of age-related decline across multiple health domains. Notably, cell type senescence signatures often emerged with unique clinical relevance that mapped to their corresponding health domain. Previously highlighted examples include the senescence burden of small airway epithelial cells best predicting COPD status (**Supp. Fig. 7a**), renal epithelial cells most strongly associating with kidney disease (**Fig. 5c**), and preadipocyte cells demonstrating the strongest association with BMI (**Fig. 5d**), among others. Importantly, the senescence signature derived from PBMCs showed strong longitudinal relevance, suggesting that not only do circulating senescence signatures shed light on current health status but can also predict future health trajectories.

Age-related decline in the immune system, or Immunosenescence, is characterized by increased susceptibility to infection and increased levels of basal inflammation, and has been previously identified as a key player in age and age-related decline.^51,52^ Additionally, Immunosenescence has been found to deleteriously impact and promote cellular senescence in multiple organs beyond the immune system,^53^ which may in part explain its role in systemic aging. Senescence of immune cells, a component of Immunosenescence, has been found to increase in prevalence with age^54^, and is also associated with worse performance across several health domains including cognitive imparment,^55^ impaired pulmonary function,^56^ and sarcopenia^57^ in human cohorts. Moreover, our group recently demonstrated that the monocyte senescence signature in circulation strongly associates with age-associated phenotypes, including measurements of overall health such as frailty and walking pace, and obesity-associated phenotypes such as blood glucose and lipids.^9^ Consistent with these reports, we observe that PBMC senescence burden, the sole indicator of immune cell senescence in the SenCat, strongly associated with clinical parameters and disease statuses in both the BLSA and InCHIANTI (**Supp. Fig. 5a-d)**. For example, the derived PBMC senescence score is most strongly associated with a frailty index as well as diabetes status in both the BLSA and InCHIANTI, consistent with similar associations with monocyte senescence we recently reported. Additionally, PBMC senescence is most strongly associated with future onset of disease including diabetes (**Fig. 6c**) and kidney disease (**Supp Fig. 8a**) in the BLSA. These findings suggest that immune cell senescence could indeed be a key player in age and disease that warrants further investigation, and that PBMC senescence markers could be particularly useful as biomarkers beyond other cell types. Most importantly, by identifying those at highest risk of future disease (diabetes, hypertension) or mortality, these senescence panels may be clinically useful to identify individuals whom to treat with senotherapeutics with the goal of reducing future risk of these outcomes. It would be of great interest to test this possibility in future senotherapeutic trials.

The findings of this study are consistent with emerging evidence suggesting that sex differences exist in senescence profiles that may contribute to sex-specific aging and health trajectories,^39^ and previous studies that have identified sex-specific differences in senescence markers in circulation between elderly males and females.^11^ There is in vitro and in vivo evidence that hormone profiles may impact senescence. For example, treatment with estrogen has been shown to reduce the onset of endothelial progenitor cell senescence in culture,^58^ and in human cohorts, hormone therapy with estrogen lowered some circulating markers of senescence in menopausal women.^59^ This study confirmed potential sex differences in the senescence burden in later life, namely in that males and females experienced peak nonlinear upregulation at different ages (**Fig. 3a**), and that these senescence markers seem to be implicated in a pro-inflammatory immune response only in males (**Fig. S2c**). This evidence, along with the finding that the senescence burden mediates the relationship between sex and diabetes status (**Fig. 3c**), suggests that this divergent senescence burden may contribute to sex-specific disease susceptibility seen in elderly populations. Importantly, lower senescence burden in both sexes was concomitant with a decrease in the prevalence of diabetes across all age groups, suggesting senotherapeutics as an intervention that should be explored in diabetes. Overall, these findings highlight that sex-specific differences in plasma senescence burden may underlie sex-specific health trajectories, a possibility that should be further studied.

Some cell type seno-age gaps showed counterintuitive clinical associations. For instance, some cell type seno-age gaps were inversely associated with disease status such as the small airway epithelial seno-age gap and obesity (**Supp. Fig. 11d**), highlighting the heterogeneity of the senescence burden. The interpretation of these results is difficult and highlights the primary limitations of this study. Although the SAPs for each cell type have been rigorously determined in two induction methods, many of these SAPs are not detected in plasma and not available for use in downstream analysis. Further, it is difficult to distinguish circulating SAPs that originate to some extent from non-senescent cell type sources in addition to senescent cell types. However, the use of signatures, rather than individual proteins, partially mitigates concerns of overlap with any given cell type. Further studies of human cohorts are needed, particularly with more plasma proteins, additional cell type signatures, longitudinal study designs, and larger cohorts, to validate the findings of the present study.

Key strengths of the study include cross-validation of most key findings in distinct aging studies. Additionally, orthogonal validation in the Human Protein Atlas, specifically the finding that SAPs exclusive to renal epithelial cells, small airway epithelial cells, PBMCs, and astrocytes generally are more highly expressed in their respective tissue, provides additional confidence that the circulating cell type-exclusive senescence burden may indeed originate in part from their respective cell type. Additionally, the demonstration that senescence signatures can predict future health trajectories is key for clinical translatability. This study is comprehensive in its identification and evaluation of both cell type-specific and core senescence signatures and provides a resource for future investigations into outcomes associated with cell type senescence burdens. Importantly, in contrast to previous efforts to identify tissue-specific protein signatures in plasma based on organ-specific RNA expression, the development of circulating senescence signatures in this study was entirely based on proteomic data originating from the SenCat.

Overall, this study comprehensively evaluates and identifies clinically relevant ‘core’ and cell type senescence signatures with cross-study validation and lays a foundation for future exploration of cell type senescence biomarkers in circulation. We demonstrate that senescence markers generally outperformed non-senescence markers in predicting clinical traits and illustrate key examples of cell type senescence signatures with unique relevance to corresponding organ systems and functions. This study highlights the potential translational use of senescence markers, particularly a high-impact panel, core, and immune cell senescence markers, that could serve as biologically relevant biomarkers and inform future clinical trials and provide higher-resolution insights into health status than previously achieved. We envision that this framework can be used to inform future senotherapeutic trials and guide the development of senotype-specific interventions.

### Limitations of the Study

The specificity of the cell type-exclusive signatures generated in this study is limited to cell types present in SenCat and cannot exclude possible contributions from other cell types. Though we have substantial evidence from both SenCat and the Human Protein Atlas that suggests that these cell type senescence markers do originate from their respective cell types and tissues, it is likely that these markers originate at least in part from other sources when tracked in vivo. Follow-up studies validating these cell type senescence markers directly in human tissues will be essential in the future. An additional limitation of this study is that a single proteomics measurement was examined from each participant; a follow-up study leveraging repeated assessments of the circulating senescence burden with time-varying cox modeling could potentially yield greater insight into associations between senescence biomarkers and health trajectories.

## RESOURCE AVAILABILITY

### Lead Contact

Requests for further information and resources should be directed to and will be fulfilled by the lead contact, Nathan Basisty.

### Materials Availability

This study did not generate new unique reagents.

### Data and Code Availability

#### Data

- All raw mass spectrometry data files and associated quantitative and statistical reports, metadata, and supplemental data are available on MassIVE (dataset identifier: MSV000096215).
- All the aggregated phenotype data for the BLSA and InCHIANTI has been provided as supplemental tables 1 and 2. To comply with patient consent and data sharing agreements, researchers are welcome and encouraged to request use of more detailed BLSA data for scientific projects by developing a pre-analysis plan that can be submitted for approval (https://blsa.nia.nih.gov/how-apply).

#### Code

- The repository of scripts used to conduct the bioinformatics analysis (version 1.0.2, DOI: 10.5281/zenodo.16237748) are available at the URL: https://github.com/geroproteomics/SenCat-Companion.^60^

### Additional Information

- Any additional information required to reanalyze the data reported in this paper is available from the lead contact upon request.

## ACKNOWLEDGEMENTS

This research was supported in part by the Intramural Research Program of the National Institutes of Health (NIH). The contributions of the NIH author(s) were made as part of their official duties as NIH federal employees, are in compliance with agency policy requirements, and are considered Works of the United States Government. However, the findings and conclusions presented in this paper are those of the author(s) and do not necessarily reflect the views of the NIH or the U.S. Department of Health and Human Services. N.B. was supported by a SenNet NIH Common Fund Grant (NIA U54 AG079779, PI: Elisseeff) and a Hevolution GRO grant (HF-GRO-23-1199068-44, PI: Basisty). BioRender was used for the generation of the Summary Figure 1.

## SUPPLEMENTAL INFORMATION

**Document S1. Figures S1–S11, Tables S1-S4, S8-S10**

**Supplemental Table 5: Protein Index for Proteins Detected in the BLSA and the SenCat**

**Supplemental Table 6: Protein Index for Proteins Detected in InCHIANTI and the SenCat**

**Supplemental Table 7: Shapiro Wilk of Distribution Differences, T-Tests Comparing SAP and Non-SAP Trait Prediction by Age Group**

**Supplemental Table 11: The top 25 selected features via elastic net modeling by cell type, trait, and study.**

**Supplemental Table 12: Nested Cross Validation Performance by Cell Type, Trait, Study, and Fold**

## STAR⍰METHODS

### KEY RESOURCES TABLE

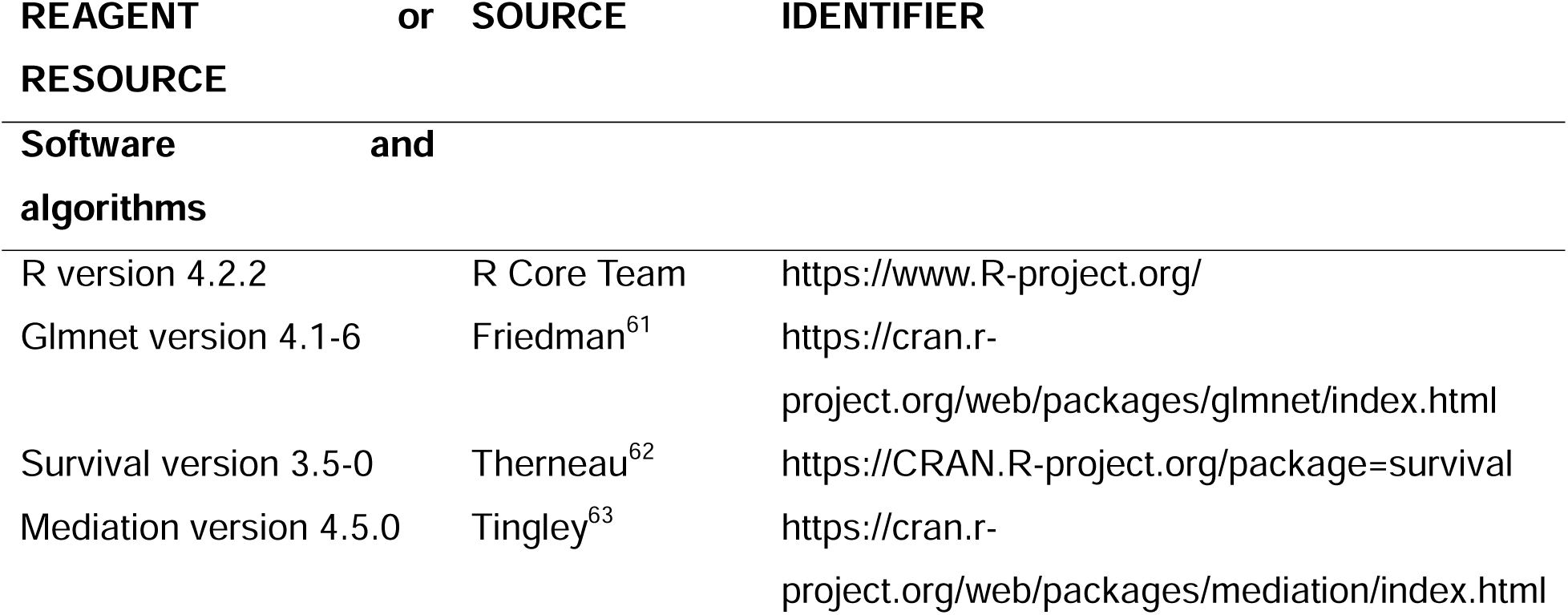

### EXPERIMENTAL MODEL AND STUDY PARTICIPANT DETAILS

#### Cell culture, senescence induction and validation

All cell types from the SenCat are of human origin, and include WI-38 lung fibroblasts (obtained from the NIGMS Human Genetic Cell Repository, Coriell Institute for Medical Research; repository ID AG06814-N), BJ skin fibroblasts (ATCC, CRL-2522), HSAEC lung epithelial cells (ATCC, PCS-301-010, HEKn epidermal skin keratinocytes (ATCC, PCS-200-010), HCAEC coronary artery endothelial cells (LifeLine Cell Technology, FC-0032), HUVEC umbilical vein endothelial cells (ATCC, PCS-100-010), HVSMC coronary artery smooth muscle cells (LifeLine Cell Technology, FC-0031), HSKM skeletal myoblasts (Gibco, A12555), PBMC peripheral blood mononuclear cells (ATCC, PCS-800-011), PreAdipo subcutaneous preadipocytes (ATCC, PCS-210-010), NHO osteoblasts (Lonza, CC-2538), NHA astrocytes (Lonza, CC-2565), HEMn melanocytes (ATCC, PCS-200-012), and HREC renal epithelial cells. Senescence induction methods include ionizing radiation, etoposide, or doxorubicin treatment. At senescence induction, all cells were at approximately 50% confluence and at low ‘population doubling’ (<5) with the exception of WI38, which were approximately at 20. Senescence induction was confirmed using BrdU incorporation, increased SA-β-gal activity, and canonical markers of senescence including GDF15, P16, P21, IL6, and LMNB1, measured using RT-qPCR analysis. For each cell type, cell pellets were processed for proteomics analysis.

#### Clinical data

Clinical data was derived from a cohort of 1275 from the Baltimore Longitudinal Study of Aging (BLSA) ages 22-96, mean age 67.52, 52.8% female. The BLSA is a long-running study of human aging that recruits subjects from the DC and Baltimore area from 1958 to present.^64^ The BLSA protocol (03AG0325) was approved by the institutional review board of the National Institute of Environmental Health Science. The second cohort of 997 examined here is from InCHIANTI, ages 21-98, mean age 66.31, 55.0% female. In both studies, sex was modeled as a covariate during statistical analysis to identify biomarkers and clinical associations irrespective of sex. InCHIANTI is a longitudinal study that recruited residents from the Chianti region of Italy and Bagno a Ripoli. The study (exemption #11976) protocol was approved by Medstar Research Institute (Baltimore, Maryland), the Italian National Institute of Research and Care of Aging Institutional Review, and the Internal Review Board of the National Institute for Environmental Health Sciences (NIEHS). For the BLSA and InCHIANTI, summary statistics as well as sample sizes for clinical parameters are provided in Tables 1 and 2, respectively.

For participants from the BLSA, walking pace refers to rapid gait speed (best time, m/s) in a physical performance test. Body composition measures were calculated using the DEXA scan method. Lean mass refers to appendicular lean mass (kg, sum of legs and arms) divided by height (km) squared. Fat percent refers to the whole body and was calculated using fat (g) divided by the sum of fat (g) and lean mass (g) multiplied by 100. Bone mineral density was calculated for the spine using the DEXA scan. Forced expiratory volume refers to the maximum air exhaled in 1 second during a pulmonary function test. Max grip refers to the maximum hand grip strength (kg) determined via hand dynamometer across multiple trials. Cognition score was calculated using the Mini-Mental State Examination. eGFR refers to estimated glomerular filtration rate calculated using the Chronic Kidney Disease Epidemiology Collaboration (CKD-EPI) equation using serum creatinine clearance, age, and sex. History of heart attack and COPD are self-report in a medical interview. Kidney Disease was assigned to those with an eGFR less than 60. Hypertension was assigned to those who self-reported, brought medication to the clinic, or had a blood pressure reading >= 130/80. Diabetes status was assigned to those who had a fasting glucose >= 126 mg/dL, a hbA1C >= 6.5%, or reported taking diabetes medication. Obesity was assigned to those with a BMI >= 30. Grip Weakness is a binary parameter assigned to those within the lowest quintile max grip strength, stratified by BMI and sex. Walk Slowness is a binary parameter assigned to those within the lowest quintile walking pace, stratified by height and sex. Additionally, a composite frailty score was created using 44 common measures of frailty, described previously,^65^ with diabetes status added as an additional component of the frailty score. Aspects of the composite score include: reported difficulty carrying out 15 basic life tasks, including walking up 10 steps, and lifting 10 pounds; self-rated health from a health status survey; five items indicating emotional state from the Center for Epidemiologic Studies Depression Scale, including feeling lonely or depressed; four items indicating cognitive status from the mini-mental state examination, including orientation to time and memory; 14 binary health conditions including cancer and cognitive impairment, among others; recent weight loss, low physical activity, walking slowness, and grip weakness.

For participants in InCHIANTI, Walking Pace refers to fast-paced walking speed across four meters (m/s). Max Grip refers to maximum grip strength in either hand measured via dynamometer. Similar to the BLSA, a Frailty Index was constructed for InCHIANTI among participants >= 65 years of age, and consisted of 5 components: weight loss not due to diet > 10 pounds in the last year; self-reported exhaustion >= 3 days in the last 7 days; self-reported low physical activity; in the 80^th^ percentile or higher for time to walk 4.57 meters; in the 20^th^ percentile or lower for grip strength. Diabetes status was determined using a fasting blood glucose of >140 (mg/dL), glycosuria, medication use, or self-report. Hypertension status was determined using blood pressure readings or self-reported status and medication. Heart Attack status was determined using self-report, documentation or ECG. COPD status was determined using evidence of either chronic bronchitis or emphysema using medication, self-report, clinical exam or documentation. Kidney function was estimated using creatinine clearance and the Cockcroft-Gault formula. Dementia status was assigned using clinical evaluation, described previously^66^, including the Mini-Mental State Examination (MMSE).

### METHOD DETAILS

#### SomaScan proteomics

The BLSA plasma proteomic study involved profiling over 7,596 SOMAmers using the 7K SomaScan Assay. The InCHIANTI study profiled over 1,322 SOMAmers using the 1.3K SOMAscan Assay at the Trans-NIH Center for Human Immunology and Autoimmunity, and Inflammation (CHI), National Institute of Allergy and Infectious Disease, National Institutes of Health (Bethesda, MD). Count normalization and performance assessment of the assay has been previously described,^67^ and reliability of SomaScan measurements have been rigorously benchmarked.^67^

#### Mass spectrometry-based proteomics

Mass spectrometry-based proteomic analysis was conducted as reported in the SenCat. Briefly, SenCat proteomic data was generated from 14 cell types, each prepared using a previously described automated SP3 protocol and acquired in data independent acquisition mode on the Orbitrap Eclipse Tribrid.^68^ Protein identification and quantification was performed in Spectronaut (v 18.1), utilizing the direct-DIA algorithm with default settings. The fasta database was derived from the UniProt human proteome (UP000005640_20191105).

### QUANTIFICATION AND STATISTICAL ANALYSIS

#### Generation of cell type-exclusive senescence signatures

Differential expression analysis was performed to identify proteins elevated in each cell type (senescent versus proliferating) using the ProtPipe package in R.^69^ The Benjamini-Hochberg method was used for multiple test correction. For each cell type within SenCat, SAPs were assigned as those with a positive log2 fold-change (sen/pro) and adjusted p-value < 0.05 in two induction methods (ionizing radiation and either etoposide or doxorubicin treatment). For the 9 cell types examined for health domain specific clinical associations, cell type-exclusive SAP were assigned only if they were a SAP in that cell type, and not a SAP in any other of the 8 cell types.

#### Elastic net modeling

All data analysis was conducted using R software^70^, version 4.2.2, in RStudio^71^. Elastic net modeling was used for feature selection using the glmnet package^61^, version 4.1-6. Elastic net is a penalized linear regression technique that selects from a group of features those that produce a model with the lowest cross-validated mean squared error. The function selects a subset of features from a provided pool using a penalty term, the strength of which is determined by the tuning parameter lambda. The technique combines two types of regularization, the stricter L1 (lasso) and less restrictive L2 (ridge). These two techniques are combined using another parameter, alpha. Elastic net modeling was performed using an alpha of 0.5, creating a mix between lasso and ridge regression to perform feature selection. For all analysis listed, only the top 25 features selected by elastic net were extracted, after ordering by effect size with the dependent variable, for each cell type and clinical parameter combination, prior to validation. For the BLSA, age, sex, and race were used as covariates during feature selection. In InCHIANTI, age and sex were used as covariates.

This study attempts to identify single senescence panels specific to individual cell types that could in the future potentially serve as biomarker panels. To achieve this, a single panel was identified using elastic net modeling on the entirety of each cohort, which were then subjected to ten rounds of cross validation (90% train, 10% test) on each cohort. While this introduces a degree of data leakage, this method is necessary to identify single panels of senescence proteins by cell type and trait. While using strict external validation, namely by performing feature selection on 90% of the data 10 times and examining each set’s predictive ability, prevents data leakage, it would produce ten unique panels of proteins for each cell type dependent on fold, which significantly complicates the identification of a potentially clinically relevant biomarker panel, and renders downstream comparisons such as relative importance (**Fig. 2c**) and mediation analysis (**Fig. 2d**) impossible. For added transparency and rigor, nested validation was performed to compare SAP vs Non-SAP trait prediction potential in both the BLSA and InCHIANTI studies, and when examining the trait prediction ability of the core senescence signature. While this does produce minor to moderate trait prediction attenuation, for many traits, T-Tests revealed that SAP proteins continued to significantly outperform the Non-SAP proteins such as with predicting age, diabetes, and hypertension, and notably, the T-Test mean difference became greater between the two groups when predicting hypertension and diabetes, and more significant when predicting diabetes (**Supp. Table 7**). These analyses independently support the conclusion that circulating SAP protein biomarkers outperform Non-SAP proteins for certain traits. The nested cross validation performance metrics by senescence status, trait, study, and fold, as well as selected features are tabulated in **Supp. Table 12**.

#### Linear, logistic regression and cox proportional hazards modeling

Pearson linear modeling and logistic regression were performed using the Stats package^70^ (Version 4.2.2), a base package of R, when identifying associations between either composite senescence scores or individual protein abundances with clinical parameters. Age, sex, and race were modeled as covariates in the BLSA using the formula: clinical parameter ∼ (protein or composite score) + age + sex + race. In InCHIANTI, age and sex were modeled as covariates using the formula: clinical parameter ∼ (protein or composite score) + age + sex. Cox proportional hazards modeling was performed using the survival package^62^ (version 3.5-0) to identify associations between either composite senescence burden scores or individual protein abundances and the onset of death or morbidities in the BLSA. To accommodate the delayed entry nature of the study, a left-truncated model was used, stratified by sex and race using the formula: Surv(Age of Proteomics Measurement, Age of event or last visit, Event Status) ∼ (protein or composite score) + strata(sex) + strata(race). When comparing SAP versus Non-SAP, or cell type senescence burden, using these modeling techniques, the senescence burden was modeled using a mean composite score of up to 25 of the elastic net selected proteins for any given cell type and clinical parameter combination. Due to variations in association, only proteins positively associated with negative traits such as frailty index and kidney disease were selected, while proteins inversely associated with positive traits such as walking pace and max grip were selected.

#### Mediation and relative importance analysis

Mediation analysis was conducted using the mediation package^63^ in R, version 4.5.0. This method was used to determine if circulating senescence proteins could mediate the relationship found between either age or sex and clinical parameters such as hypertension or diabetes. Elastic Net Modeling was used to identify senescence signatures implicated in each clinical parameter, limited to the top 25 by effect size per group. These proteins were condensed into a composite senescence burden score by taking the z-scored mean abundance per participant. The mediation package determines the proportion of the association between sex or age and clinical parameters that is mediated by the composite senescence burden score. Covariates included in the model were sex, race, and the alternate composite score. A 95% confidence interval was determined using bootstrapping with 1000 iterations. Similarly, the SAP and Non-SAP composite scores were compared for their relative importance using the relaimpo package,^72^ version 2.2-7, identifying the proportion of the variation of clinical parameters explained using each score while controlling for the other score, with 95% confidence intervals calculated using bootstrapping with 1000 iterations.

#### Pathway analysis

Gene ontology was represented via overrepresentation analysis using the ClusterProfiler package, version 4.6.0.^73^ Biological process was the ontology class selected for all ontology analysis. For ontology figures (**Supp. Fig. S1c-d**) the universe supplied consisted of all proteins detected in both the SenCat and the BLSA 7k SomaScan. All displayed output of overrepresentation analysis was statistically significant with Benjamini-Hochberg multiple test correction.

