## Supplementary Figures and Supplementary Tables 1, 2, 3, 4, 8, 9, 10 for "Circulating Cell Type Senescence Signatures Reveal High-Resolution Health Status and Trajectories in Human Longitudinal Studies"

Supplemental Figure 1: Comparing Senescence and Non-Senescence Signatures for Trait Prediction

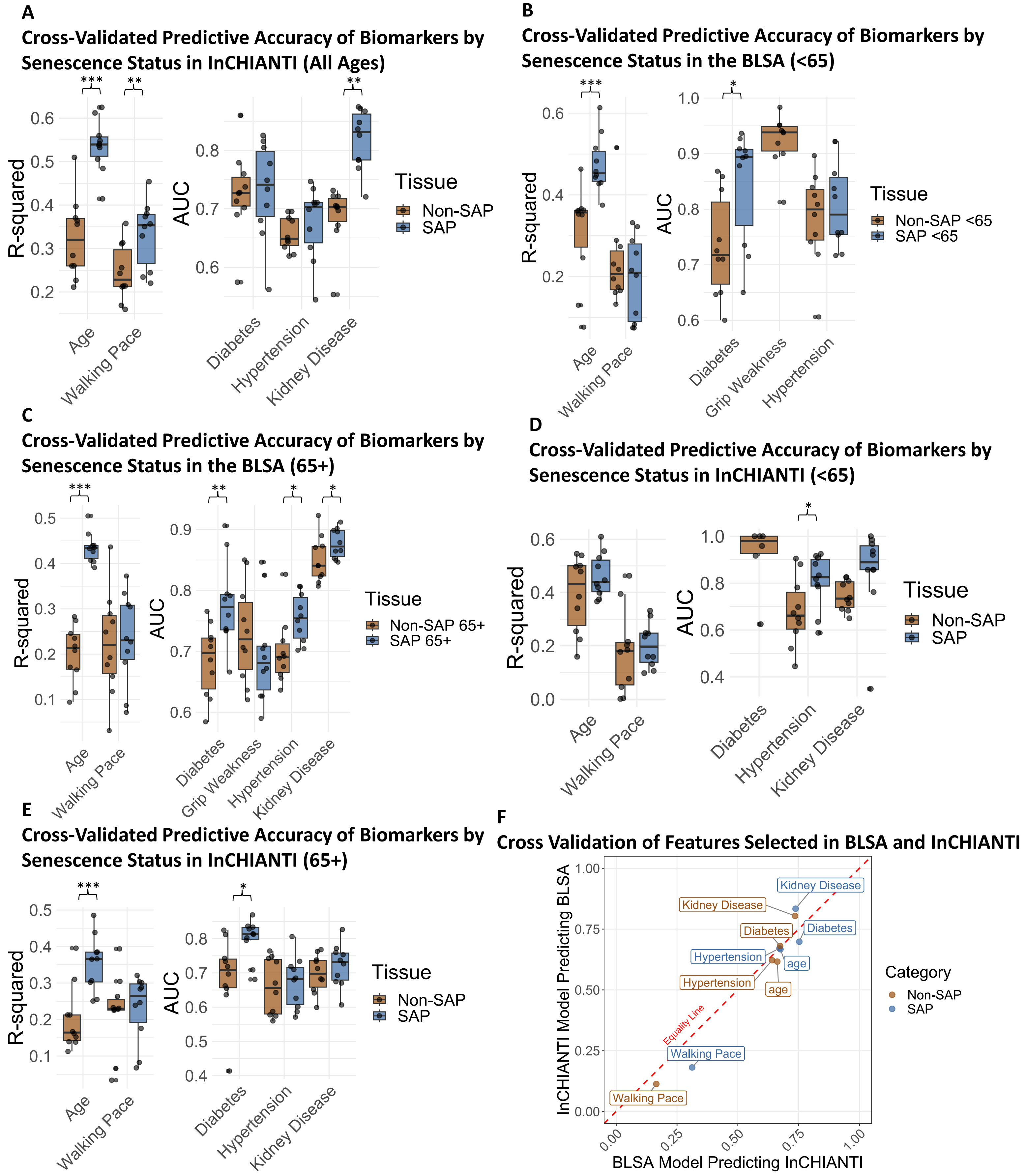

Supp. Figure 1: Cross-validated R-Squared values across ten iterations (90% train, 10% test) for clinical parameters. To normalize different pool sizes, only the top 25 ENSPs were used for each group of proteins, which were condensed into a mean composite score. **A)** SAP and Non-SAP senescence burden score trait prediction in the InCHIANTI. **B)** SAP and Non-SAP senescence burden score trait prediction in the BLSA among participants younger than 65. **C)** SAP and Non-SAP senescence burden score trait prediction in the BLSA among participants 65 years of age or older. **D)** SAP and Non-SAP senescence burden score trait prediction in InCHIANTI among participants younger than 65. **E)** SAP and Non-SAP senescence burden score trait prediction in the InCHIANTI among participants 65 years of age or older.

Supplemental Figure 2: Stratified Random Sampling of the BLSA and Ontology of Proteins Upregulated in Later Life

**A**

**Age Distribution of Total BLSA Cohort, Eligible Participants (Ages 50-90), and Those Randomly Selected**

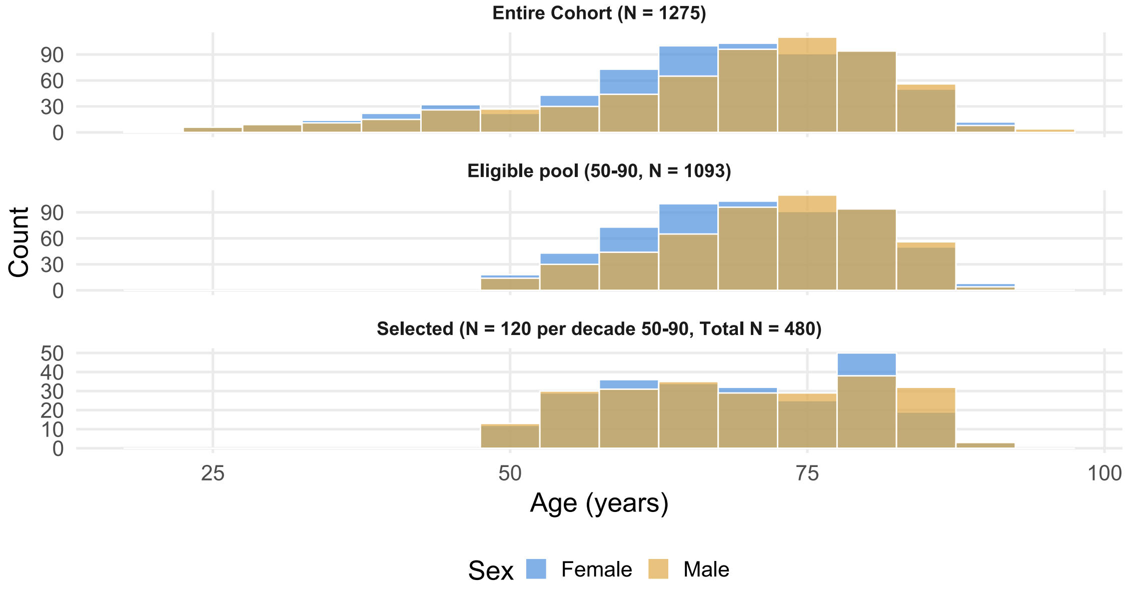

**B**

**Overrepresentation Analysis of SAPs by Stage of Life**

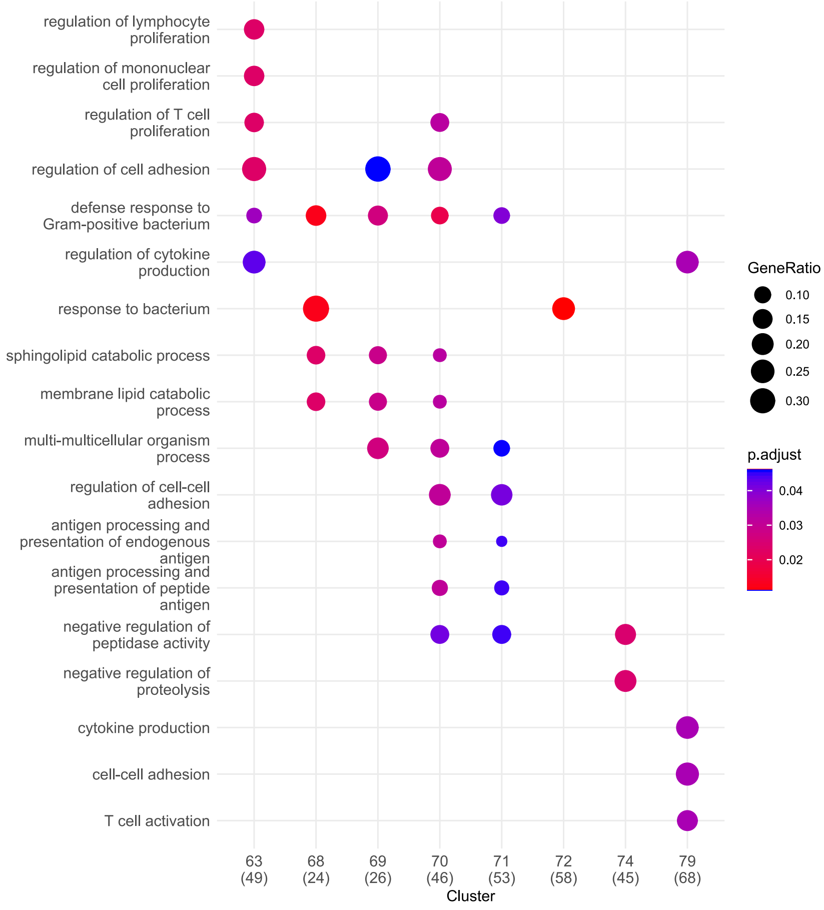

**C**

**Overrepresentation Analysis of SAPs by Sex**

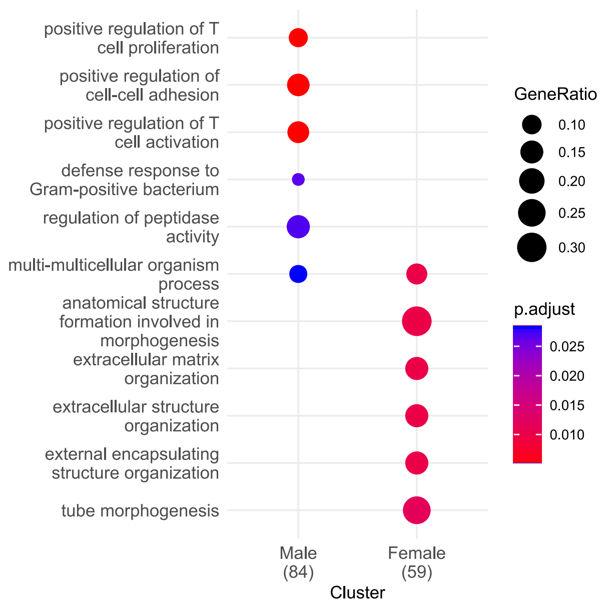

Supp. Figure 2: Differential Expression Sliding Window Analysis (DE-SWAN) was performed to identify SAPS and Non-SAPs upregulated by year from ages 60-80. **A)** Age distribution of the BLSA entire cohort, the eligible pool of individuals ages 50-90, and those randomly selected, N = 120 per decade, 50% male and 50% female. **B)** Overrepresentation analysis (Biological Process) of SAPs that are elevated by year according to the DE-SWAN method in the BLSA. **C)** Overrepresentation analysis (Biological Process) of SAPs that are elevated in males or females during the 60s or 70s according to the DE-SWAN method in the BLSA.

#### Supplemental Figure 3: Senescence Markers Identified by Tissue and Induction Method

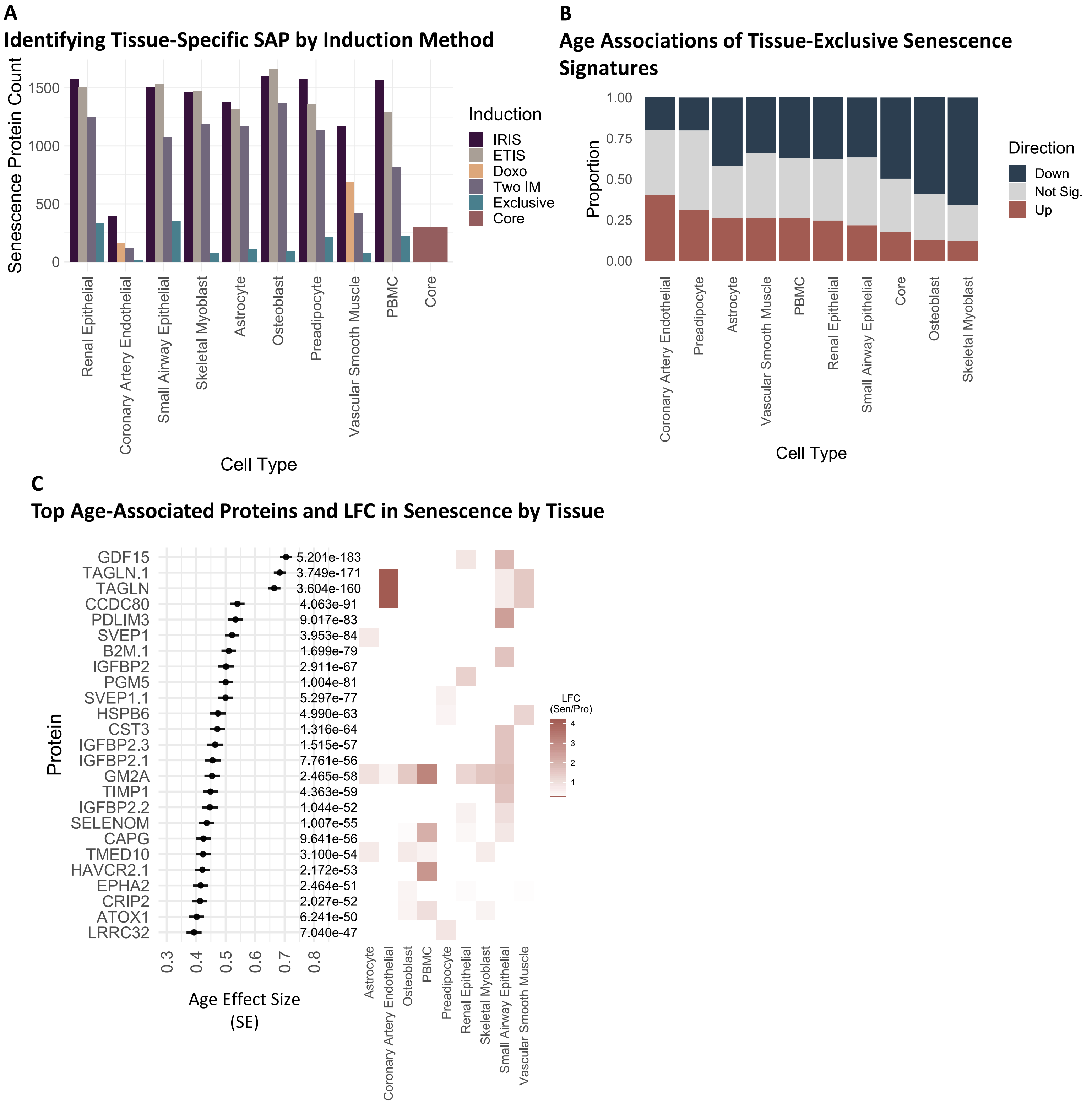

Supp. Figure 3: A) Senescence associated proteins (SAPs) were identified across cell types with multiple induction methods, including ionizing radiation (IRIS), etoposide (ETIS), and doxorubicin (DOXO). SAPs present in two induction methods (Two IM), as well as those present in two induction methods and exclusive to each cell type (Exclusive), were also identified by cell type. Last, a group of proteins that were elevated in two induction methods in at least 6 cell types were identified as a core senescence signature (Core). B) Linear modeling of cell type-exclusive SAPs revealed the proportion of each that were associated with age in the BLSA using the formula: age ~ protein + sex + race. C) The top 25 age-associated SAPs among all cell types in the SenCat by their effect size and adjusted p-value for age, and their respective average log fold change (Sen/Pro) in each tissue.

*For Sen/Pro log fold change, only proteins significantly upregulated in that cell type using two induction methods are shown by their average LFC across both induction methods.

### Supplemental Figure 4: Orthogonal Validation of Cell Type SAPs in the Human Protein Atlas

**A**

#### Orthogonal Validation of the SenCat Cell Type Senescence Signatures in the Human Protein Atlas

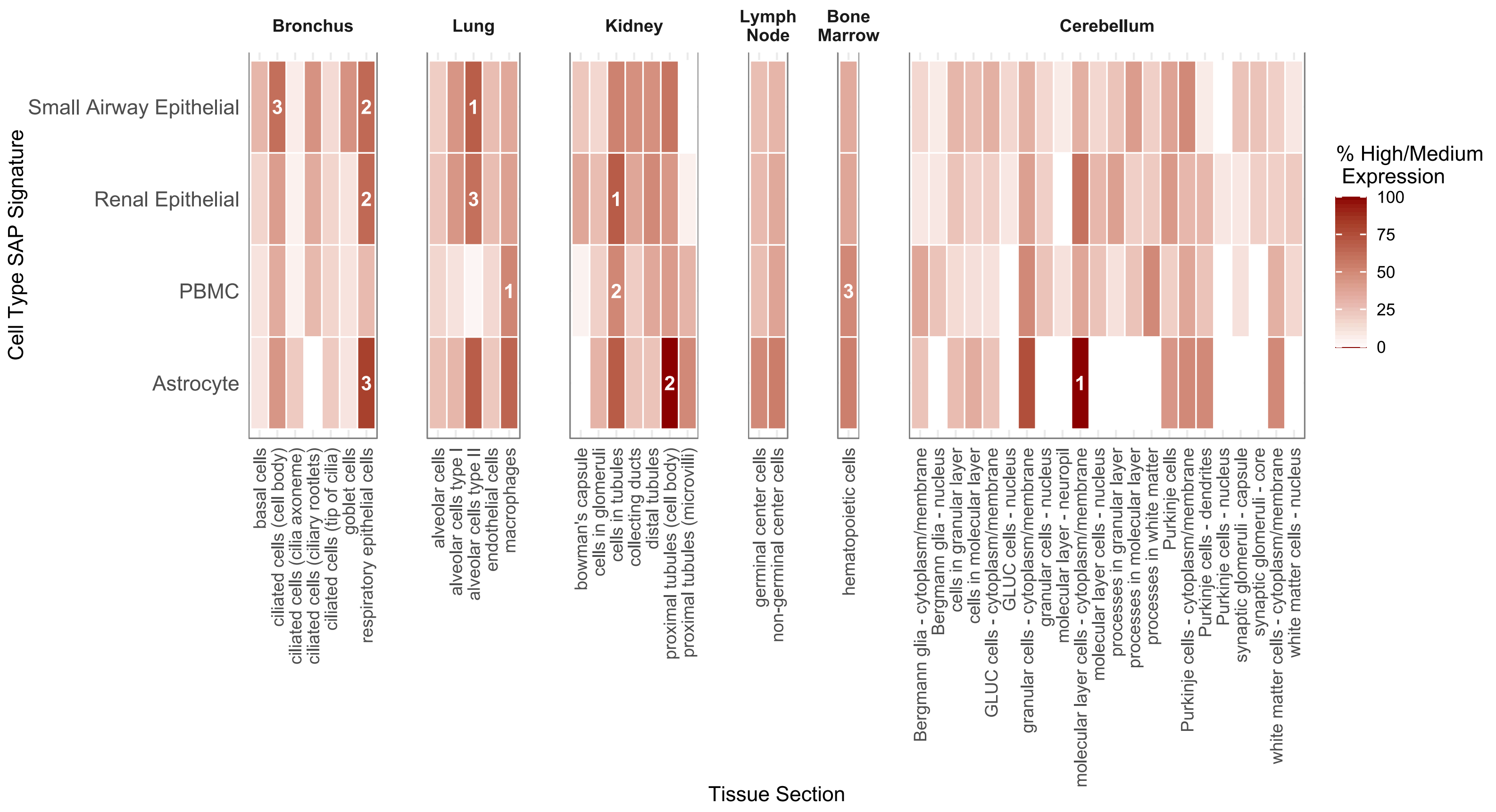

Supp. Figure 4: A) Four cell type enriched senescence signatures, including from small airway epithelial cells, renal epithelial cells, PBMCs, and astrocytes, were examined in the Human Protein Atlas in tissues that roughly correspond to the same organ system. The percent of the cell type SAPs for each signature with medium or high expression compared with no or low expression is shown. Numbers 1, 2, and 3 represent the three tissue sections where the highest percentage of medium or highly expressed proteins from each given cell type senescence signature was observed.

### Supplemental Figure 5: Senescence Burden Associations with Clinical Parameters

## A
#### BLSA Senescence Burden Associations with Continuous Traits

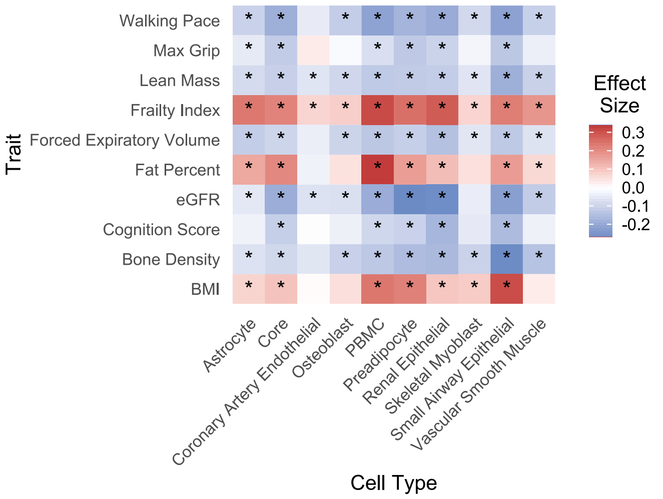

## B
#### BLSA Senescence Burden Associations with Disease States

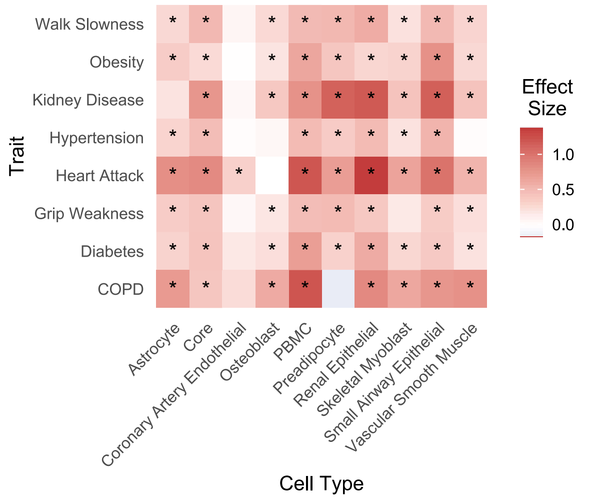

## C
#### InCHIANTI Senescence Burden Associations with Continuous Traits

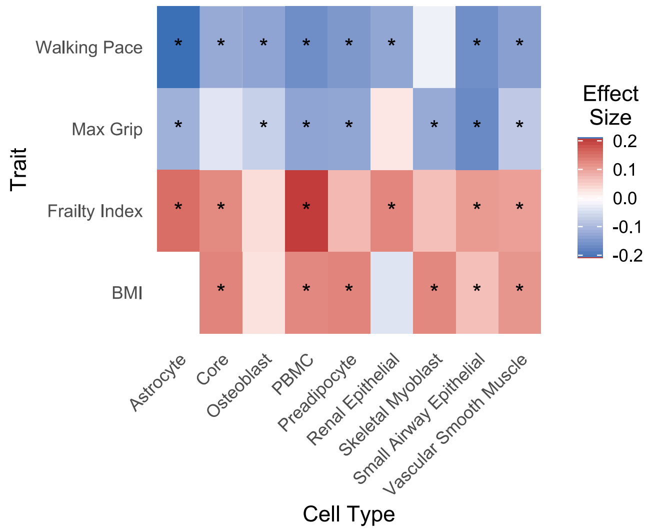

## D
#### InCHIANTI Senescence Burden with Disease States

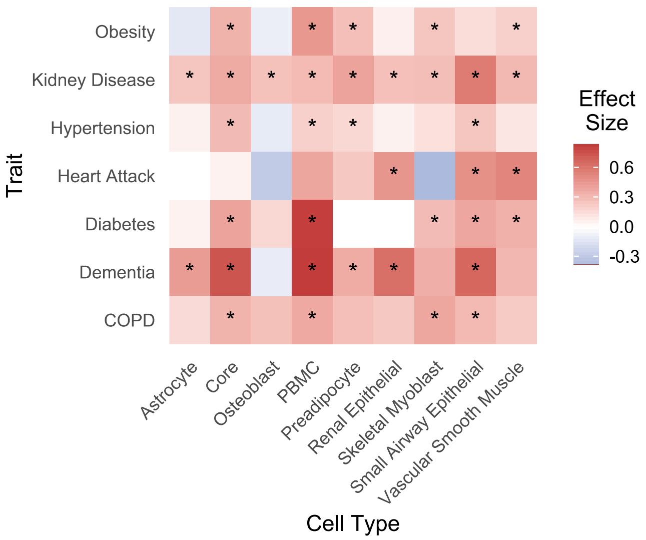

Supp. Figure 5: Up to 25 cell type-exclusive SAPs were selected using elastic net modeling for each cell type and clinical parameter combination. Each set was compiled into a mean composite score to represent cell type senescence burden implicated in clinical parameters. A) Unbiased display of cell type composite senescence burden associations via linear modeling with continuous clinical traits in the BLSA. B) Cell type composite Senescence Burden associations via logistic regression with disease status in the BLSA. C) Cell type composite Senescence Burden associations via linear modeling with continuous clinical traits in InCHIANTI. D) Cell type composite Senescence Burden associations via logistic regression with disease status in InCHIANTI.

### Supplemental Figure 6: Comparing Senescence and Non-Senescence Signatures for Trait Prediction

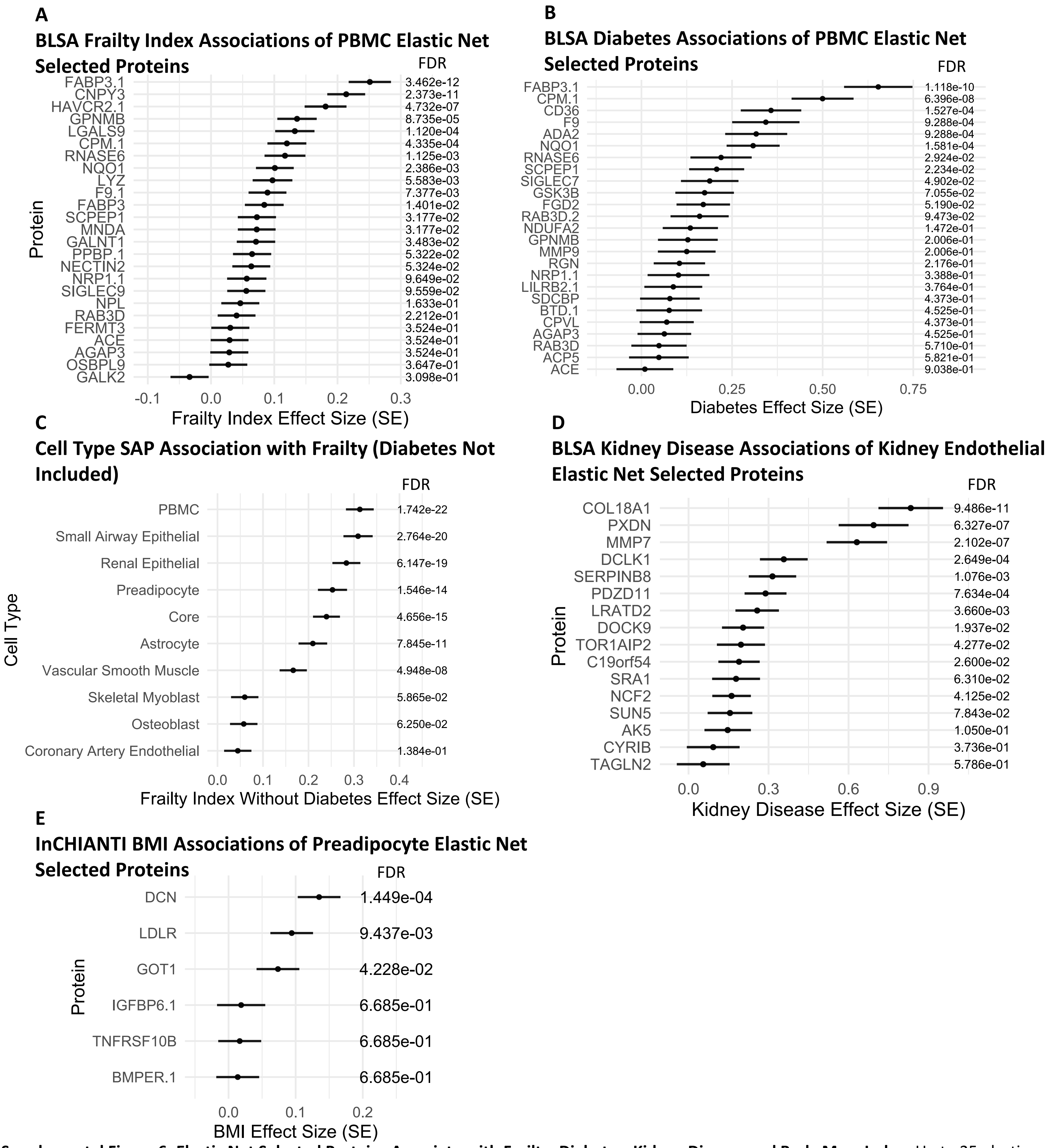

**Supplemental Figure 6: Elastic Net Selected Proteins Associate with Frailty, Diabetes, Kidney Disease and Body Mass Index.** Up to 25 elastic net selected SAPs were selected from cell type-unique senescence signatures as implicated in a frailty index or separately for diabetes status, kidney disease, or BMI which were each compiled into a mean composite score to represent cell type-unique senescence burden. A) Frailty index associations via linear modeling for PBMC SAPs selected via elastic net modeling in the BLSA using the formula: frailty index ~ protein + age + sex + race. B) Diabetes associations via logistic regression for PBMC SAPs selected via elastic net modeling in the BLSA using the formula: diabetes ~ protein + age + sex + race. C) Cell Type SAP composite score associations with a 43-component Frailty score (Diabetes Excluded). D) Kidney disease status associations with renal epithelial SAPs selected via elastic net modeling in the BLSA using logistic regression with the formula: kidney disease status ~ protein + age + sex + race. E) BMI associations via linear modeling for preadipocyte SAPS selected via elastic net modeling in InCHIANTI using the formula: BMI ~ protein + age + sex.

Supplemental Figure 7: Tissue-Unique Senescence Signatures Show Unique Clinical Predictive Capacity

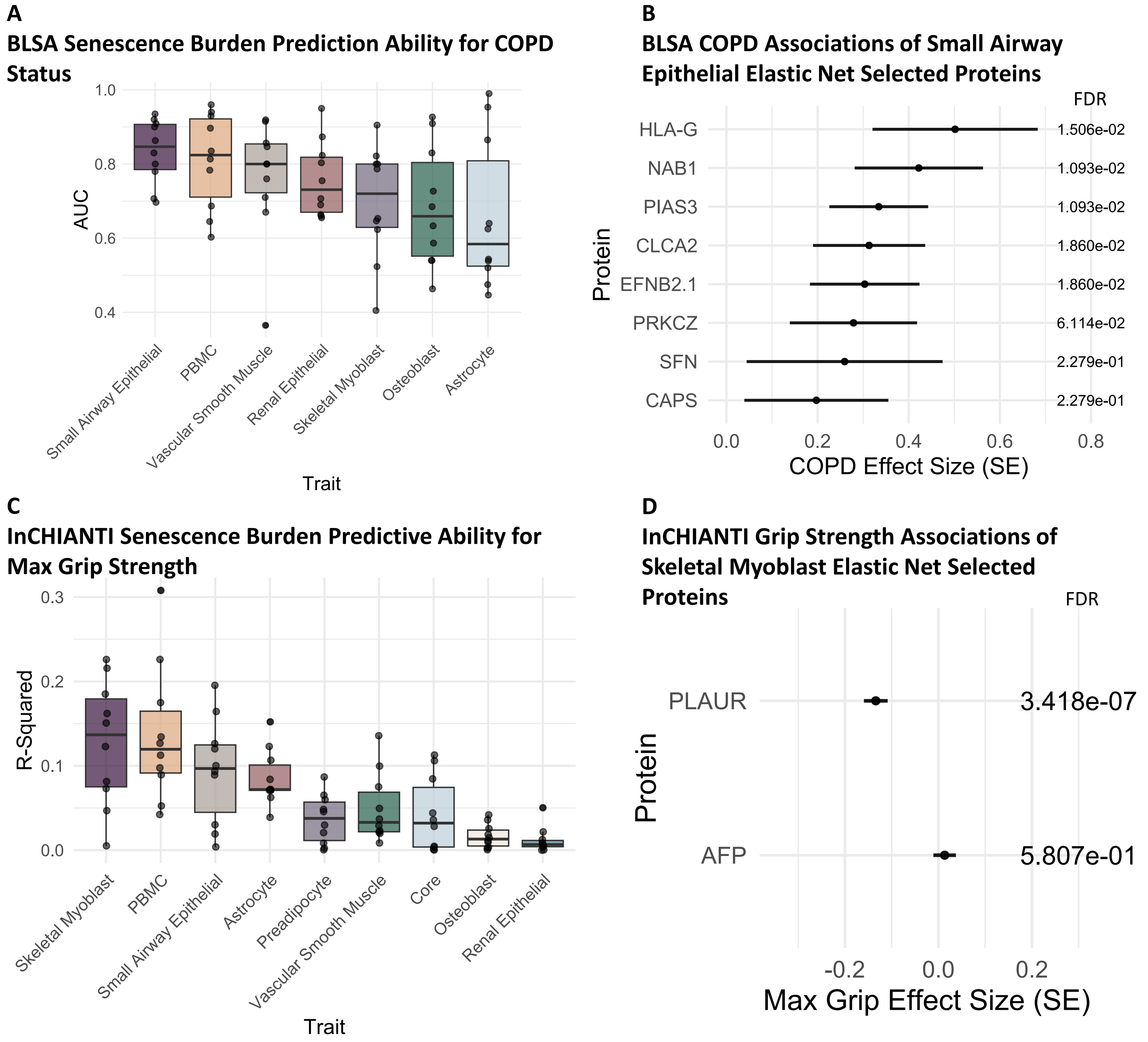

Supp. Figure 7: Up to 25 cell type-exclusive SAPs were selected via elastic net modeling across cell types and were each compiled into a mean composite score to represent cell type senescence burden implicated in clinical parameters. A) The cross-validated predictive ability of cell type senescence burden and COPD in the BLSA. B) COPD associations via logistic regression for Small Airway Epithelial SAPs selected via elastic net modeling in the BLSA. C) The cross-validated predictive ability of cell type senescence burden and Max Grip Strength in InCHIANTI. D) Max Grip Strength associations via linear modeling for Skeletal Myoblast SAPs selected via elastic net modeling in InCHIANTI.

Supplemental Figure 8: Tissue-Specific Senescence Burden Reveals Longitudinal Health Trajectories

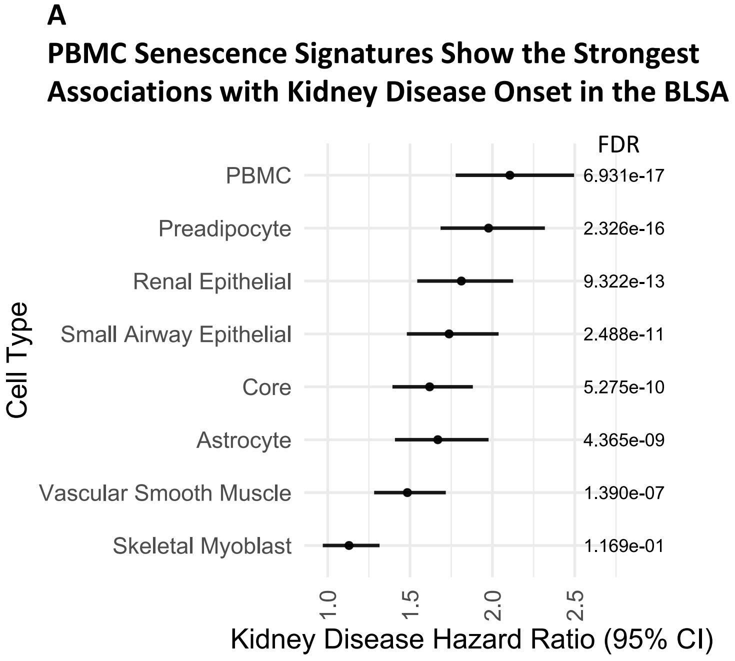

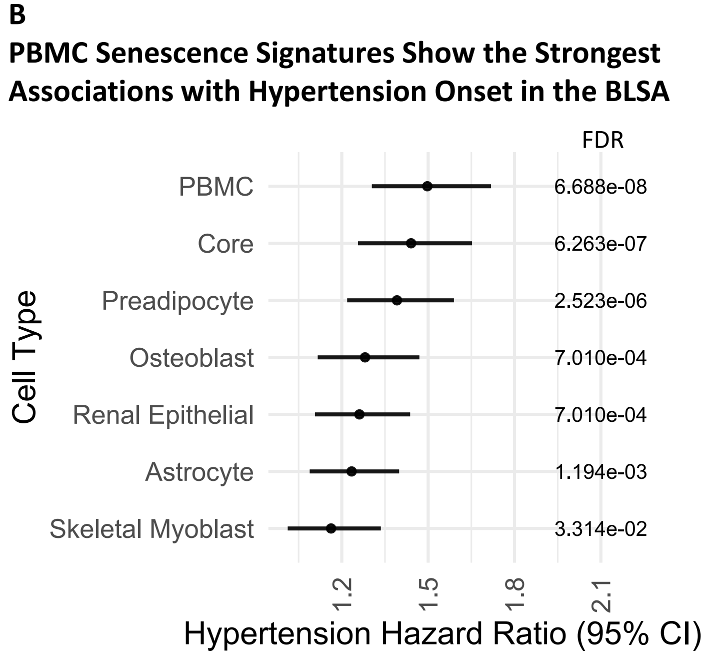

Supp. Figure 8: Up to 25 cell type-exclusive SAPS were selected via elastic net modeling and each set was compiled into a mean composite score to represent cell type senescence burden across multiple cell types. **A**) The association between the cell type senescence burden and kidney disease onset via cox proportional hazards modeling in the BLSA. **B**) The association between the cell type senescence burden and hypertension onset via cox proportional hazards modeling in the BLSA.

Supplemental Figure 9: Tissue-Specific Seno-Age Shows Different Trajectories

**A**

**Predicted Seno-Age Show Unique Lifespan Trajectories - BLSA**

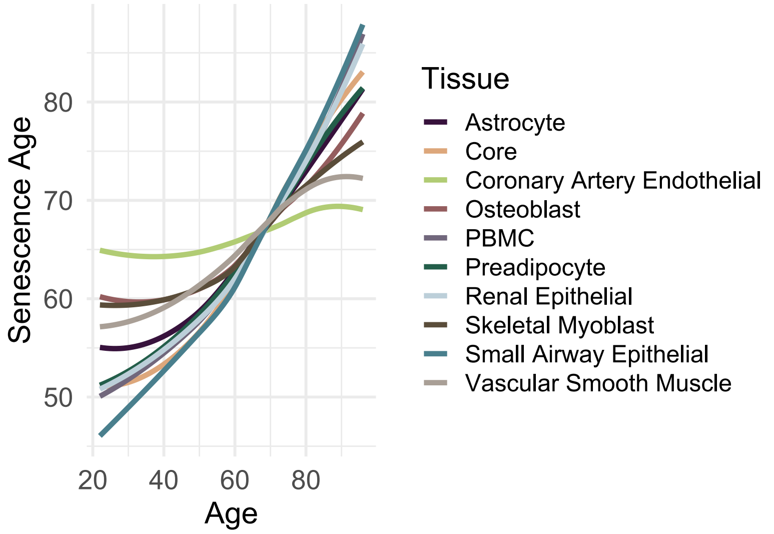

**B**

**Predicted Seno-Age Show Unique Lifespan Trajectories - Inchianti**

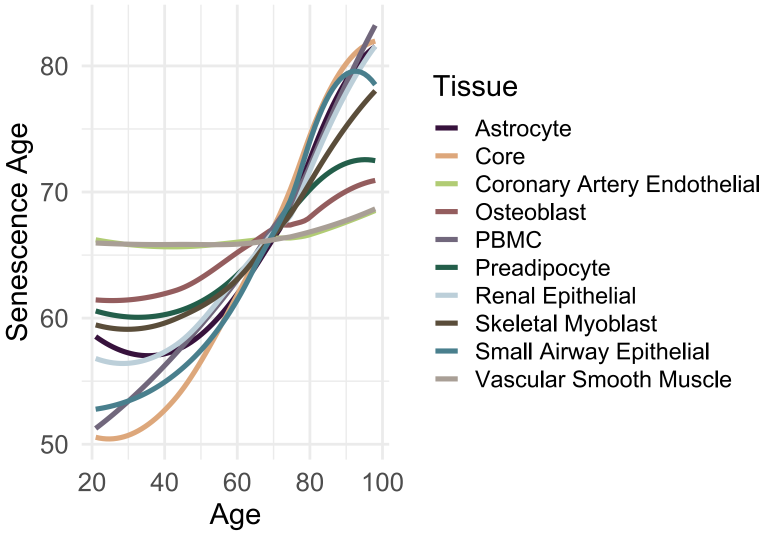

**C**

**Seno-Age Correlations - BLSA**

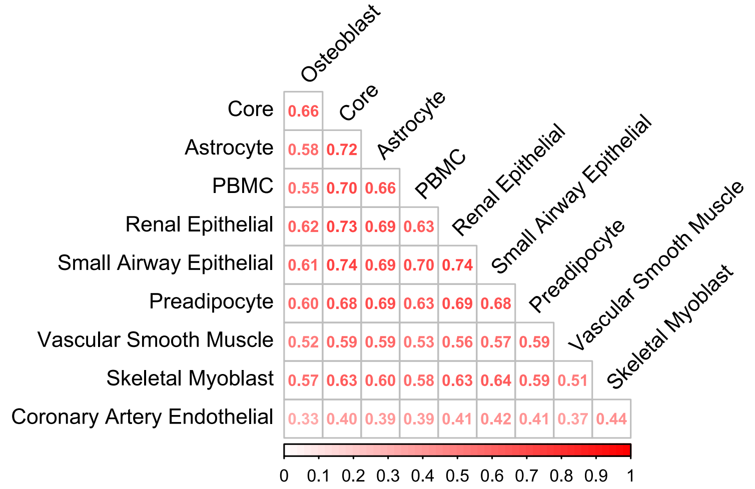

**D**

**Seno-Age Correlations - Inchianti**

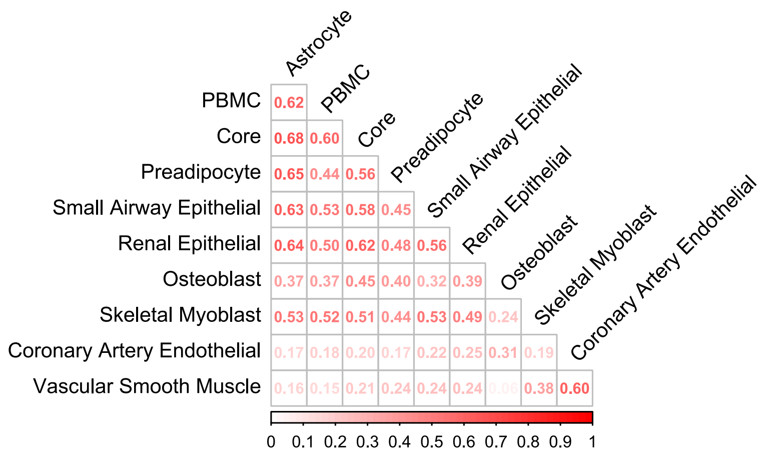

Supp. Figure 9: A) Up to 25 cell type-exclusive SAPs were selected via elastic net modeling to predict age for each cell type. These ENSPs were used to model age using linear modeling for each SenCat cell type, termed Seno-Age. Seno-age trajectories are shown by cell type and chronological age in the BLSA. B) As in A, seno-age is shown by cell type and chronological age in InCHIANTI. C) Seno-Age spearman correlations are shown by cell type in the BLSA D) As in C, Seno-Age spearman correlations are shown by cell type in Inchianti.

Supplemental Figure 10: Deriving Senescence Age and Senescence Age Gaps in the BLSA

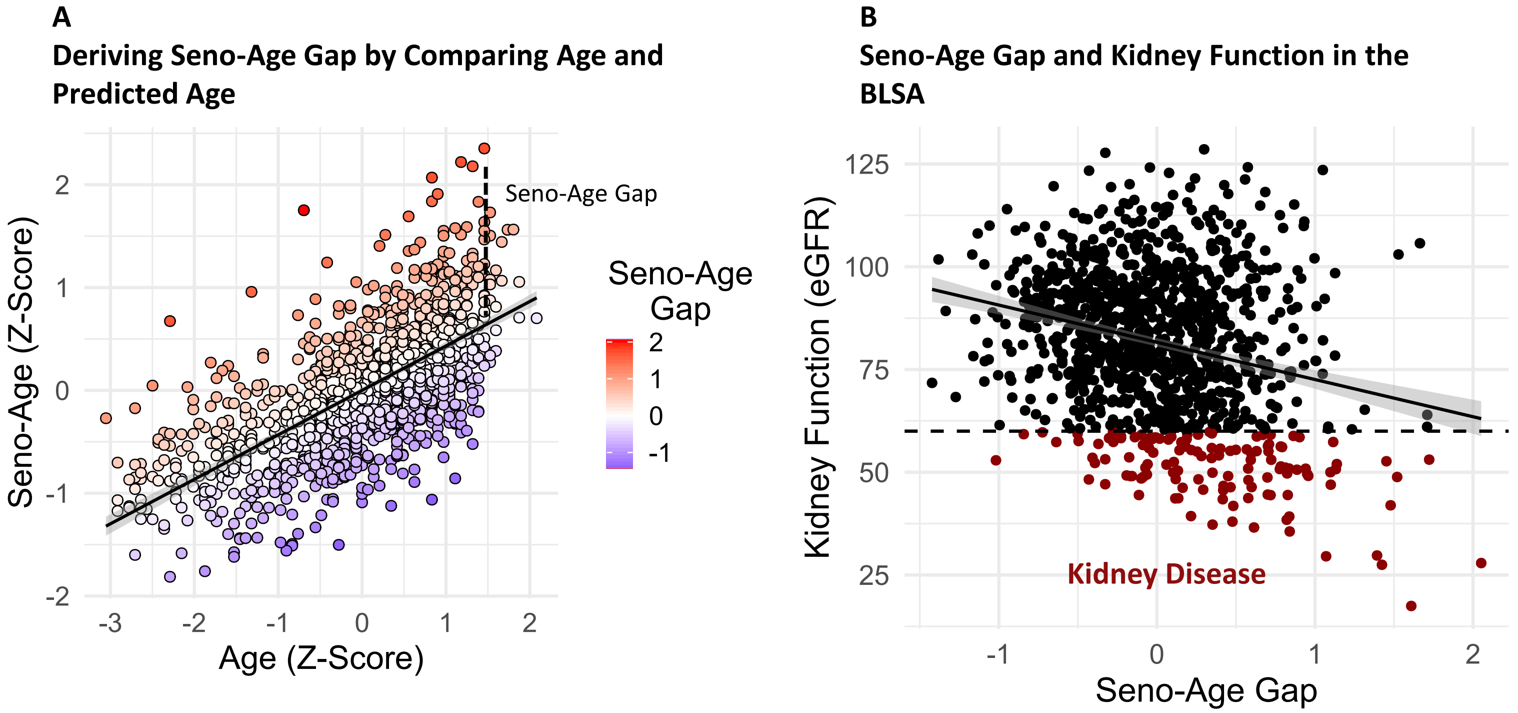

Supp. Figure 10: Up to 25 cell-type exclusive SAPs were selected using elastic modeling for each cell type. Each set was used to predict age using a linear model for each individual, representing "seno-age" for each cell type. Seno-age gaps were created using the residual of a linear trend line between chronological age and modeled seno-age. **A**) Example seno-age shown is predicted using astrocyte elastic net selected SAPs and compared with chronological age. **B**) Example of seno-age gap compared with kidney function in the BLSA. The seno-age gap shown was determined using astrocyte elastic net selected SAPs.

#### Supplemental Figure 11: Seno-Age Gap Associations with Clinical Parameters

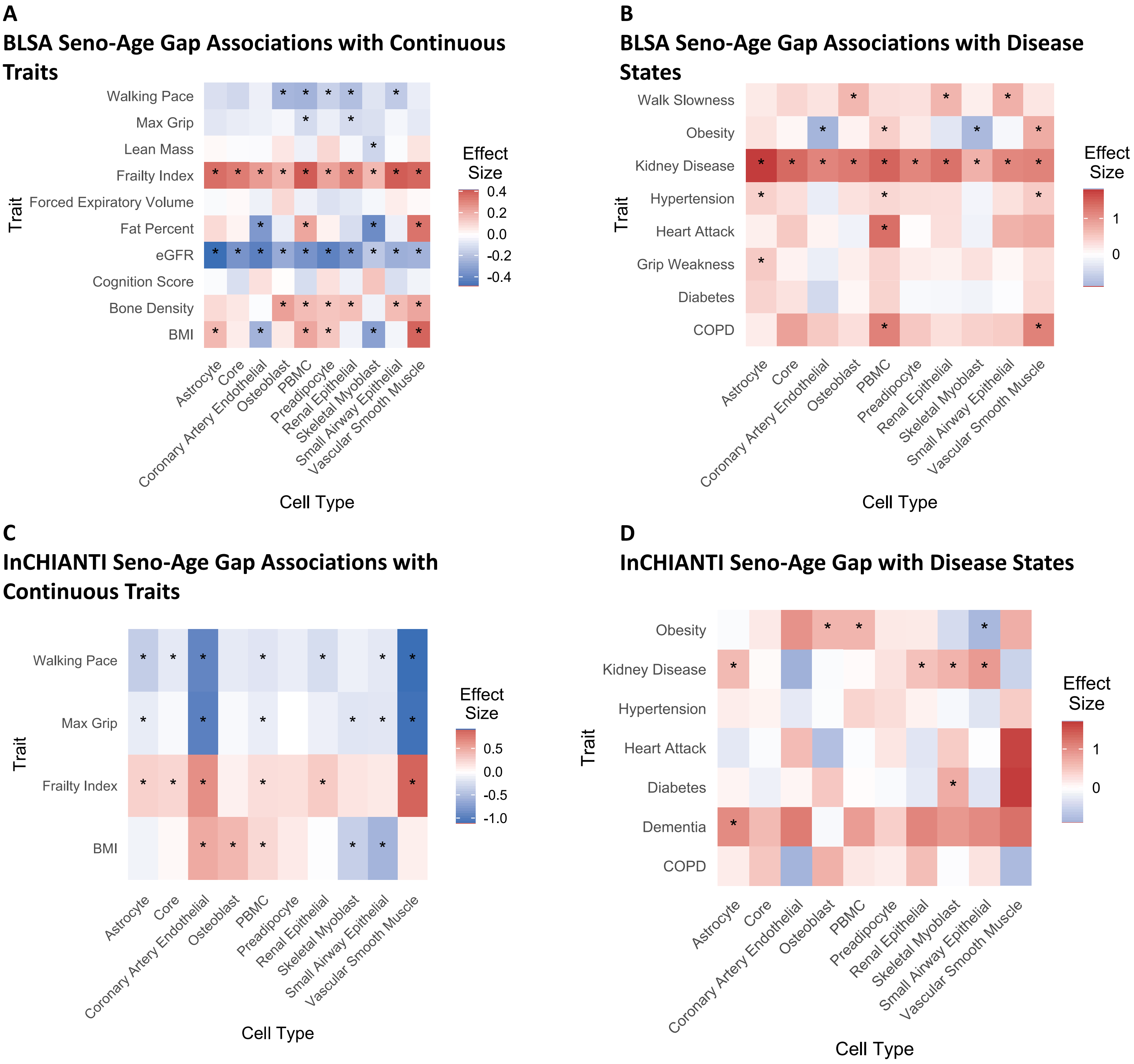

Supp. Figure 11: Up to 25 cell type-exclusive SAPs selected via elastic net modeling were used to model age within a linear model across multiple cell types, termed cell type seno-age. From here, cell type seno-age gaps were produced using the residuals of a trend line between seno-age and chronological age. A) Unbiased display of Seno-Age Gap associations via linear modeling with continuous clinical parameters in the BLSA. B) Seno-age gap associations via logistic regression with disease status in the BLSA. C) Seno-Age Gap associations via linear modeling with continuous clinical parameters in InCHIANTI. D) Seno-Age Gap associations via logistic regression with disease status in InCHIANTI.

Supplemental Table 1: Senescence induction dose by cell type.

| Cell Type | Senescence trigger | Treatment details | Days into senescence |
|---|---|---|---|
| HCAEC (Arterial Endothelial) | Doxo | 250 nM, 3 days | 8 |
| HCAEC (Arterial Endothelial) | IR | 15 Gy | 8 |
| HUVEC (Venal Endothelial) | Etoposide | 10 uM, 3 days | 8 |
| HUVEC (Venal Endothelial) | IR | 7.5 Gy | 8 |
| HSAEC (Lung Epithelial) | Etoposide | 20 uM, 3 days | 8 |
| HSAEC (Lung Epithelial) | IR | 10 Gy | 8 |
| BJ (Skin Fibroblasts) | Etoposide | 25 uM, 6 days (refreshed at day 3) | 8 |
| BJ (Skin Fibroblasts) | IR | 15 Gy | 8 |
| WI-38 (Lung Fibroblasts) | Etoposide | 50 uM, 6 days (refreshed at day 3) | 8 |
| WI-38 (Lung Fibroblasts) | IR | 15 Gy | 8 |
| HSKM (Myoblasts) | IR | 10 Gy | 8 |
| HSKM (Myoblasts) | Etoposide | 20 uM | 8 |
| PreAdipo (Subcutaneous Preadipocytes) | IR | 15 Gy | 8 |
| PreAdipo (Subcutaneous Preadipocytes) | Etoposide | 25 uM | 8 |
| HVSMC (Vascular Smooth Muscle) | Doxo | 85 nM, 3 days | 8 |
| HVSMC (Vascular Smooth Muscle) | IR | 15 Gy | 8 |
| HEKn (Skin Keratinocytes) | Etoposide | 25 uM, 3 days | 8 |
| HEKn (Skin Keratinocytes) | IR | 10 Gy | 8 |
| HEMn (Melanocytes) | Etoposide | 25 uM, 3 days | 8 |
| HEMn (Melanocytes) | IR | 7.5 Gy | 8 |
| NHO (Osteoblasts) | Etoposide | 20 uM, 3 days | 8 |
| NHO (Osteoblasts) | IR | 10Gy | 8 |
| NHA (Astrocytes) | Etoposide | 20 uM, 3 days | 8 |
| NHA (Astrocytes) | IR | 10Gy | 8 |
| PBMC (Peripheral Blood Mononuclear) | Etoposide | 20 uM, 3 days | 6 |
| PBMC (Peripheral Blood Mononuclear) | IR | 7.5 Gy | 6 |
| HREC (Renal Mixed) | Etoposide | 20 uM, 3 days | 8 |
| HREC (Renal Mixed) | IR | 10 Gy | 8 |

Supplemental Table 2: Clinical Parameters in the BLSA - Data reported as mean (standard deviation).

| | All | 20 to 40 | 40 to 60 | 60 to 70 | 80+ | N |
|---|---|---|---|---|---|---|
| Count | 1275 | 70 | 252 | 702 | 251 | |
| Age | 67 (14.1) | 32.5 (4.6) | 51.2 (6) | 70.2 (5.4) | 83.4 (3.2) | |
| Female Percent | 52.8 | 54.3 | 56.3 | 52.3 | 50.2 | |
| BMI | 26.9 (4.6) | 25.1 (4.8) | 27.1 (4.8) | 27.5 (4.8) | 25.6 (3.3) | 1269 |
| Walking Pace | 1.8 (0.4) | 2.1 (0.3) | 2 (0.3) | 1.8 (0.3) | 1.6 (0.3) | 983 |
| eGFR | 81.6 (17.6) | 107.2 (12.6) | 93.3 (13.6) | 77.9 (15.1) | 69.8 (13.6) | 1120 |
| Bone Density | 1.1 (0.2) | 1.1 (0.1) | 1.1 (0.2) | 1.1 (0.2) | 1.1 (0.2) | 1022 |
| Lean Mass | 7.5 (1.3) | 7.9 (1.3) | 7.9 (1.4) | 7.4 (1.3) | 7.1 (1.1) | 1021 |
| Fat Percent | 35.3 (9.4) | 29.5 (10.1) | 34.3 (9.3) | 37 (9.2) | 33.6 (8.5) | 1021 |
| Forced Expiratory Volume | 2.5 (0.9) | 3.5 (0.9) | 3 (0.9) | 2.4 (0.8) | 2 (0.7) | 1001 |
| Max Grip | 33.5 (11.4) | 40.7 (12) | 38.9 (11.7) | 32.2 (10.1) | 26.8 (9.2) | 1019 |
| Frailty Index | 0.1 (0.1) | 0 (0) | 0.1 (0) | 0.1 (0.1) | 0.1 (0.1) | 980 |
| Cognition Score | 28.5 (1.6) | 29.3 (0.9) | 29.2 (1) | 28.5 (1.7) | 28.1 (1.7) | 1047 |

Supplemental Table 3: Clinical Parameters in InCHIANTI - Data reported as mean (standard deviation).

| | All | 20 to 40 | 40 to 60 | 60 to 70 | 80+ | N |
|---|---|---|---|---|---|---|
| Count | 997 | 99 | 104 | 665 | 129 | |
| Age | 66.3 (15.4) | 30.3 (5.2) | 49.5 (5.8) | 70.7 (4.5) | 84.9 (3.6) | |
| Female Percent | 55 | 54.5 | 49 | 54.3 | 63.6 | |
| BMI | 27.2 (4.1) | 24.3 (3.6) | 27.2 (4) | 27.8 (4.1) | 26.1 (3.9) | 962 |
| Walking Pace | 1.5 (0.4) | 1.9 (0.3) | 1.8 (0.3) | 1.5 (0.3) | 1.1 (0.4) | 934 |
| Max Grip | 33.1 (13.8) | 46.2 (13.5) | 43.3 (14.4) | 31.7 (12) | 21.8 (8.5) | 843 |
| Frailty Index | 1.6 (0.8) | NaN (NA) | NaN (NA) | 1.5 (0.7) | 1.9 (0.9) | 334 |

Supplemental Table 4: Counts and Proportions of SAP and Non-SAP in the BLSA and InCHIANTI Studies

| | | | |
|---|---|---|---|
| Raw Numbers | | | |
| | SAP | Non-SAP | Inconclusive |
| BLSA | 1941 | 1266 | 737 |
| InCHIANTI | 355 | 228 | 129 |
| Fisher's Exact Test P-Value: 0.9332 | | | |
| Proportions | | | |
| | SAP | Non-SAP | Inconclusive |
| BLSA | 49.21399594 | 32.09939148 | 18.68661258 |
| InCHIANTI | 49.85955056 | 32.02247191 | 18.11797753 |

Supplemental Table 8: Features Selected in Both BLSA and InCHIANTI Studies for Predicting Traits

| Trait | BLSA | InCHIANTI | ModelType | Type | Features |
|---|---|---|---|---|---|
| age | 0.685414306 | 0.6729173 | Linear | SAP | Q99988/Q76M96/P00750/Q9GZN4/Q9UBP4.1/Q12841/P24593.1/P46531/Q9UM47/O15031.1/Q12860/P52799/P40121/P00747.2/Q8N423/P22897/P35442.1/P05155.1/P17931/P09758.1/Q99538/P35613/P45985/P15260/Q03154/Q10588/O95881.1/P08758/P28799/P42892/P20936 |
| Walking Pace | 0.311743502 | 0.180708363 | Linear | SAP | P05413.1/Q76M96/P09237/P17931/Q01469/Q99988/Q13813 |
| Hypertension | 0.669854722 | 0.671115417 | Logistic | SAP | P05413.1/Q99988/O14786.1/P00734.1/P09237/Q8TDQ0.1/Q9Y286/Q10588/Q08380 |
| Diabetes | 0.75122549 | 0.69670351 | Logistic | SAP | Q99988/O15031.1/O00764/P16671/P06396.1/P18065.2 |
| Kidney Disease | 0.73802823 | 0.834049077 | Logistic | SAP | P61769.1/Q96HD1/Q01973.1/P39060 |
| age | 0.664525355 | 0.617492387 | Linear | Non-SAP | Q99988/Q76M96/P00750/Q9GZN4/Q9UBP4.1/Q12841/P24593.1/P46531/Q9UM47/O15031.1/Q12860/P52799/P40121/P00747.2/Q8N423/P22897/P35442.1/P05155.1/P17931/P09758.1/Q99538/P35613/P45985/P15260/Q03154/Q10588/O95881.1/P08758/P28799/P42892/P20936 |
| Walking Pace | 0.16515581 | 0.112268806 | Linear | Non-SAP | P05413.1/Q76M96/P09237/P17931/Q01469/Q99988/Q13813 |
| Hypertension | 0.640447942 | 0.622111567 | Logistic | Non-SAP | P05413.1/Q99988/O14786.1/P00734.1/P09237/Q8TDQ0.1/Q9Y286/Q10588/Q08380 |
| Diabetes | 0.672690918 | 0.680749467 | Logistic | Non-SAP | Q99988/O15031.1/O00764/P16671/P06396.1/P18065.2 |
| Kidney Disease | 0.734198832 | 0.803495021 | Logistic | Non-SAP | P61769.1/Q96HD1/Q01973.1/P39060 |

Supplemental Table 9: Standardized Mean Differences between a group included in DE-SWAN and an excluded group.

| | | | |
|---|---|---|---|
| Trait | Selected Group | Removed Group | SMD |
| Age | 69.3 (10.7) | 72.2 (7.9) | -0.312 |
| Female | 50.00% | 44.50% | 0.11 |
| BMI | 27.4 (4.6) | 26.9 (4.5) | 0.114 |
| Walking Pace | 1.8 (0.4) | 1.8 (0.3) | 0.087 |
| Diabetes | 14.20% | 18.20% | -0.107 |
| Hypertension | 49.40% | 48.90% | 0.009 |

Supplemental Table 10: Validating Proportional Hazards Assumptiom of SAPs Implicated in Mortality

| Protein | chisq | pval | symbol |
|---|---|---|---|
| P09104.1 | 0.094544687 | 0.758477132 | ENO2 |
| P55058 | 0.049958195 | 0.823136034 | PLTP |
| O00391 | 1.990745366 | 0.158262967 | QSOX1 |
| P30085.1 | 1.325517206 | 0.24960428 | CMPK1 |
| Q9H1C4 | 0.211836516 | 0.645331754 | UNC93B1 |
| Q9UBI4 | 1.254626748 | 0.26267063 | STOML1 |
| Q9UKA8 | 4.186452811 | 0.040748273 | RCAN3 |
| Q8NFI3 | 0.033245735 | 0.855320516 | ENGASE |
| P09237 | 7.60555959 | 0.00581886 | MMP7 |
| P42785 | 5.449954648 | 0.019568835 | PRCP |
| Q13162 | 2.914742697 | 0.087773411 | PRDX4 |
| P35442.1 | 7.518723445 | 0.006106094 | THBS2 |
| P05120 | 1.160230622 | 0.281417717 | SERPINB2 |
| Q8NBJ7 | 6.888008952 | 0.008677587 | SUMF2 |
